# Detection and Continuous Monitoring of Memory Dysfunction in Mild Cognitive Impairment and Healthy Aging Through Adaptive Computational Phenotyping

**DOI:** 10.1101/2024.03.15.24304345

**Authors:** Holly S. Hake, Maarten van der Velde, Thomas J. Grabowski, Hedderik van Rijn, Andrea Stocco

**Affiliations:** Department of Psychology, University of Washington; Seattle, 98195, USA; Graduate Program in Neuroscience, University of Washington; Seattle, 98195, USA; Precision Cognition Labs; Groningen, Netherlands; Department of Experimental Psychology, University of Groningen; Groningen, Netherlands; Behavioral and Cognitive Neurosciences, University of Groningen; Groningen, Netherlands; Departments of Radiology and Neurology, University of Washington; Seattle, 98195, USA

**Keywords:** Memory, Memory Decline, Memory Assessment, Mild Cognitive Impairment, Computational Model, Computational Phenotyping

## Abstract

With the rising prevalence of age-related memory impairments, efficiently detecting and monitoring decline is increasingly urgent. Unfortunately, traditional assessment methods fall short of these needs, as they typically require in-person administration and cannot be repeated frequently. Here, we demonstrate that remote identification and monitoring of abnormal memory function is possible by combining an online assessment platform with computational phenotyping, allowing repeatable, unsupervised remote observations from patients. Fifty-one well-characterized older individuals, including 24 patients with amnestic mild cognitive impairment and 27 age- and education-matched healthy controls, completed a series of longitudinal, unsupervised, remote weekly 8-minute online memory assessments for up to one year. Weekly test data were fit to a formal model of memory consolidation and forgetting, yielding an individualized index of memory function, the Seattle-Groningen Memory Assessment (SGMA) score. The SGMA score was found to be reliable, with a mean correlation of *r* = 0.70 across assessments. The score was also found to be stable across different study materials, and only barely affected by practice effects, which averaged to a 0.2% increase per assessment. Finally, the SGMA score was found to be diagnostic, being capable of detecting mild cognitive impairment with up to 87% accuracy. These findings show that model-based, adaptive assessments can support high-frequency, remote detection and scalable longitudinal monitoring of early memory decline, providing a new way to assess memory decline trajectories in healthy aging and dementia.

**Author Summary:** Memory decline is a central feature of aging and an early marker of neurodegenerative disease, yet current methods for assessing memory are poorly suited for frequent, long-term monitoring. Standard neuropsychological tests are time-consuming, administered in person, and cannot be repeated often without substantial practice effects. As a result, changes in memory function are often detected too late and their trajectories cannot be traced.

In this study, we show that memory function can be reliably assessed and monitored remotely using a brief, unsupervised online task combined with computational modeling. Older adults, including individuals with mild cognitive impairment, completed short weekly memory assessments from home over periods of up to one year. Rather than relying on raw test scores, we used a digital twin approach: each participant’s data was fitted to a mathematical model of memory consolidation, and its forgetting parameter was used to derive a precise measure of memory function, the Seattle–Groningen Memory Assessment (SGMA) score.

We find that this model-based score is reliable over time, largely unaffected by repeated testing, and able to distinguish individuals with mild cognitive impairment from healthy older adults with high accuracy. These results demonstrate that digital, model-driven cognitive assessments can support scalable, high-frequency monitoring of memory decline. Such approaches may complement existing clinical tools and enable earlier detection and more precise tracking of cognitive changes in aging and dementia.

## Introduction

Age-related memory impairments, such as those caused by Alzheimer’s disease and related degenerative disorders, pose a significant public health challenge. Because some etiologies of memory impairment respond to early treatment (*1*), timely detection is critical. Detection, however, depends on the availability of reliable and repeatable assessments. Unfortunately, current tools are often time-consuming, culturally biased, and require administration by trained specialists. Hwang et al (*2*) evaluated various neuropsychological tests for detecting dementia and mild cognitive impairment (MCI) and found that their detection accuracy varied widely, ranging from 60 to 90% across studies; because of this variability none of the tests could be recommended in isolation. Additionally, traditional neuropsychological tests are prone to practice effects or retest effects (*3*), where performance improves over consecutive tests even without a corresponding cognitive amelioration. This can lead to the underdiagnosis of MCI and similar conditions (*4–6*) and restricts the ability to do the high-frequency repeated testing necessary for accurately tracking cognitive changes over shorter time spans. Finally, the lack of adaptivity of traditional neuropsychological tests requires participants to respond to all items, including ones that will be too easy or too difficult for them. This makes traditional assessments unnecessarily burdensome and two to 20 times longer than adaptive ones (*7*). Compounded, these limitations contribute to growing health disparities, as diagnostic services are more accessible in urban and wealthier areas compared to rural and lower-income regions, and might contribute to underdiagnosis rates of up to 75% for Alzheimer’s disease (AD) (*8*).

Access to memory assessments can be expanded through the online administration of neuropsychological tests (*9*, *10*). However, their increased public availability and discussion (*11*) have led to overexposure, making some tests overly familiar and further reducing their effectiveness. This issue is serious enough that the American Academy of Clinical Neuropsychology issued a statement on “test security,” (*12*) urging that neuropsychological tests not be publicly disseminated to preserve their validity. Thus, even online versions of these assessments remain ill-suited for monitoring progressive disease and response to treatment. In addition, as highlighted by Ratcliff and McKoon (*13*), it is often unclear what the traditional neuropsychological tests measure. While tests are claimed to measure a specific construct, the mapping from test to construct remains ambiguous. Ratcliff and McKoon therefore propose a shift toward *computational phenotyping*—a theory-driven approach that uses computational models of cognitive processes to better understand how those processes differ between cognitively unimpaired individuals and those with cognitive disorders (*13–15*).

The present study demonstrates, for the first time, the viability of this approach in the domain of memory impairments. By using a detailed model of memory consolidation and forgetting, we achieved accurate and reliable computational phenotyping of early-stage memory impairments such as MCI on a remote, mobile platform in a longitudinal study. Specifically, we used an existing adaptive online declarative-knowledge learning system based on a well-established memory model (*16–18*) to create an unsupervised, repeatable, and remote assessment system. This system tailors the presentation of stimuli based on an online estimate of the patient’s forgetting rate. By adapting the sequence of stimuli to the memory capacities of individual users, it significantly reduces the burden of the assessment protocol while maintaining high accuracy and reliability.

The underlying model (Fig 1A), is based on four key principles: (1) memories are created through the accumulation of episodic traces (*19*); (2) forgetting occurs as the activation that is associated with each trace fades over time according to a power law (*20*); (3) the activation of a memory is a function of the summed activation of all traces (*16*); (4) the activation of a memory determines both the probability of its retrieval and, together with estimates for the latency associated with non-memory processes, the time required for its retrieval (*21*).

**Fig 1.**
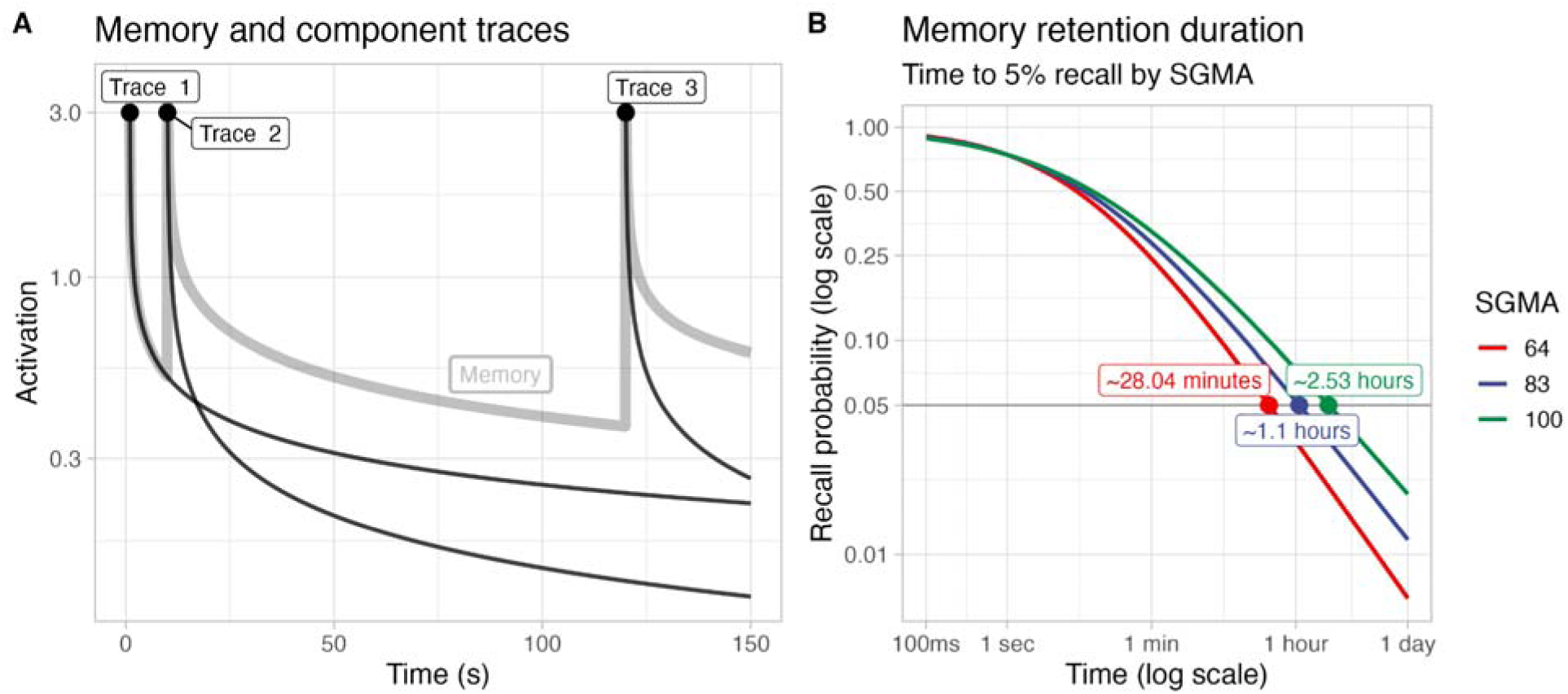
Computational model of memory activation and recall probability. **(A)**Simulated activation decay of a memory (gray line) composed of three memory traces (black lines) encoded at different time points (0, 10, and 120 seconds). Each trace follows its own decay rate, with earlier traces decaying slower, while later traces that build upon a higher activation decay more rapidly. **(B)** Relationship between SGMA score and memory retention, showing recall probability over time on a log scale. Higher SGMA scores are associated with longer retention, as indicated by the time when recall probability drops below 5% for three hypothetical individuals with SGMA scores of 100 (typical score for cognitively unimpaired young adults), 83 (score of the cognitively unimpaired participants in this study), and 64 (score of participants diagnosed with MCI).

The speed at which a memory’s first episodic trace fades over time is a trace-specific parameter, referred to as the Speed of Forgetting (SoF). Subsequent presentations of the same information result in the creation of new traces, whose fading rate depends on both the initial SoF and the memory’s activation at the time of their creation (*17*). With any presentation, the system’s software can infer a memory’s activation from the user’s accuracy and response time, and thereafter adjust the original SoF to reduce the mismatch between predicted and observed behavior (*21*). An interpretable, individual assessment of memory function, called the Seattle-Groningen Memory Assessment (SGMA) score, is then generated as a normalized average of all trace-specific SoF values. Because of its algorithmic nature, the SGMA score can be used to make realistic behavioral predictions; for example, it can be used to estimate the probability that an individual would remember a given fact when quizzed at different intervals, or how long an individual can hold on to a newly encoded fact before forgetting it (Fig 1B).

This approach enables an individualized and rapid assessment of memory performance that is characteristic of an individual (*22–24*) and reflects a combination of underlying biological processes (*25*), some of which are affected by aging and compromised in degenerative dementia.

To examine the validity of our computational phenotyping method, we conducted a longitudinal study of cognitively unimpaired elderly adults and elderly individuals with MCI. We chose individuals with MCI, rather than dementia, to both ensure participants had sufficient cognitive abilities to perform the task over the duration of the study and to test the model’s ability to detect earlier, more subtle differences in memory function (*15*, *26*). This cohort of individuals was followed for a maximum of 12 months, during which they performed weekly online assessments with our model-based system to characterize their SGMA scores. We hypothesized that (1) SGMA scores would be reliable across repeated assessments; (2) individuals with MCI would exhibit lower SGMA scores than cognitively unimpaired controls, with accelerated forgetting observed over time; (3) SGMA scores would have clinical validity, allowing for the reliable early identification of abnormal memory function.

## Methods

### Participants

Fifty-one participants were recruited on a rolling basis from the University of Washington Alzheimer’s Disease Research Center. All participants provided informed consent and were compensated for their participation in the online memory game portion of the study. All recruitment and testing procedures were conducted in accordance with the Declaration of Helsinki and approved by the University of Washington’s Institutional Review Board.

The inclusion criteria for the study were as follows: (1) age between 55 and 85 years, (2) fluency in English, and (3) no major medical or psychiatric conditions that would affect cognitive performance. Participants were classified into two groups: those with mild cognitive impairment (MCI; *N* = 24; 5 female aged 63-76, 19 male aged 66-83) and cognitively unimpaired controls (*N* = 27; 19 female aged 56-84, 8 male aged 59-77). Participants were selected to ensure that both groups were similar in terms of age (controls: 69.2 ± 4.9; MCI: 72.8 ± 7.2) and years of education (MCI: 16.8 ± 2.5, controls: 17.6 ± 1.9); it was not possible to balance sex. Fig 2 provides an overview of two group characteristics.

**Fig 2.**
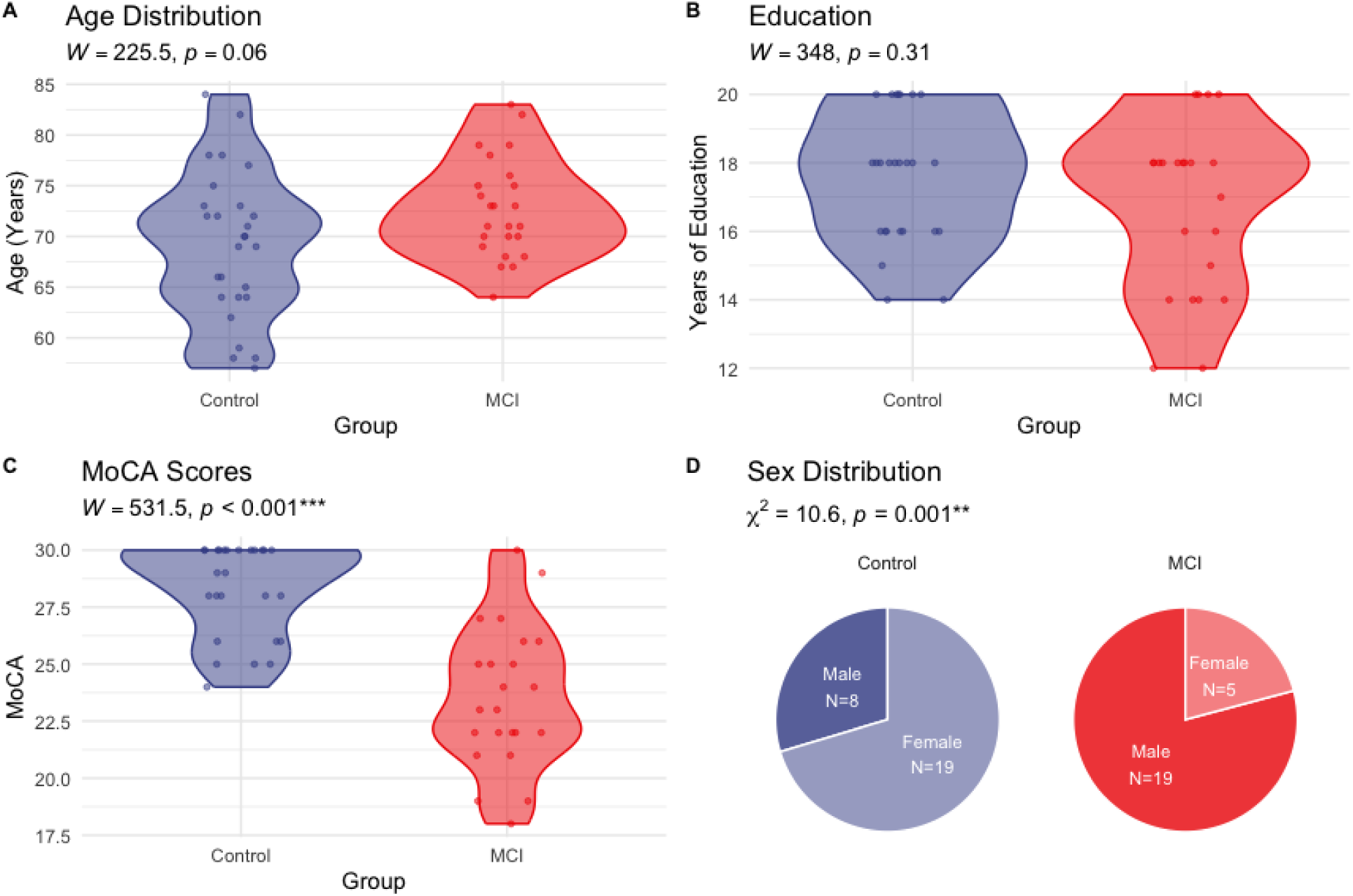
A comparison of the MCI and Control group characteristics. **(A)** The age distribution of individual participants was not significantly different between MCIs (*M* = 72.7, *SD* = 4.9) and controls (*M* = 69.2, *SD* = 7.2; Wilcoxon *W* = 225.5, *p* = 0.06). **(B)** The distribution of years of education for individual participants was also not significantly different between MCIs (*M* = 16.8, *SD =* 2.5) and controls (*M* = 17.6, *SD* = 1.9: Wilcoxon *W* = 348, *p* = 0.31). **(C)** MoCA scores for individual participants, showing significant lower scores in the MCI group (*M* = 23.6, *SD* = 3.0) than in the control group (*M* = 28.3, *SD* = 2.0; Wilcoxon *W* = 531, *p* < 0.001). **(D)** Sex composition across both groups, showing higher number of males in the MCI group (*N* = 19) than in the control group (*N* = 8) \*\*\**p* < 0.001, ** *p* < 0.01, * *p* < 0.05

### Patient Selection and Diagnosis

MCI was diagnosed using a combination of methods including clinical evaluation, cognitive testing, and medical history. The clinical evaluation was conducted by a geriatric psychiatrist or a neurologist, who assessed the participant’s cognitive and functional abilities using standardized tools. Cognitive testing was performed using a battery of neuropsychological tests that measured various cognitive domains such as memory, attention, and executive function. Medical history was obtained through a structured interview and review of medical records. Participants were classified as having MCI if they had a Clinical Dementia Rating scale ≤ 0.5. Controls were screened for cognitive impairment using the same methods as MCI participants. They were classified as cognitively unimpaired if they scored within normal limits on cognitive tests and had no history or reports of cognitive decline or functional impairment.

All participants with memory impairment were classified as having amnestic MCI at the moment of enrollment. During the study, one participant was reclassified as having non-amnestic MCI, one was diagnosed with Alzheimer’s dementia upon re-assessment, and one was diagnosed as having Autism Spectrum Disorder in addition to memory impairments. Of the 21 individuals who were classified as amnestic MCI, eight exhibited deficits only in the memory domain, and 13 exhibited deficits in multiple domains, such as executive function, language, or visual reasoning (*27*).

### Adaptive Memory Assessment

Weekly at-home assessments were completed with the online adaptive learning system described in Sense et al. (*22*) and hosted by MemoryLab.nl, allowing participants to engage remotely with a computer, tablet, or smartphone. The system implements a retrieval practice paradigm (*28*) by presenting new study pairs (e.g., “France / Paris”) and scheduling repeated tests (e.g., “France / ?”) at strategic points based on the online estimates of a user’s SoF. Fig 3 provides an example of the software interface. Test probes included only the cue of a pair, e.g., “France / ?”; users responded by selecting one of four multiple-choice options on a screen. The sessions, which participants referred to as “lessons” or “memory games”, covered a range of topics including nature, history, art, science, geography, media, food, and culture.

**Fig 3.**
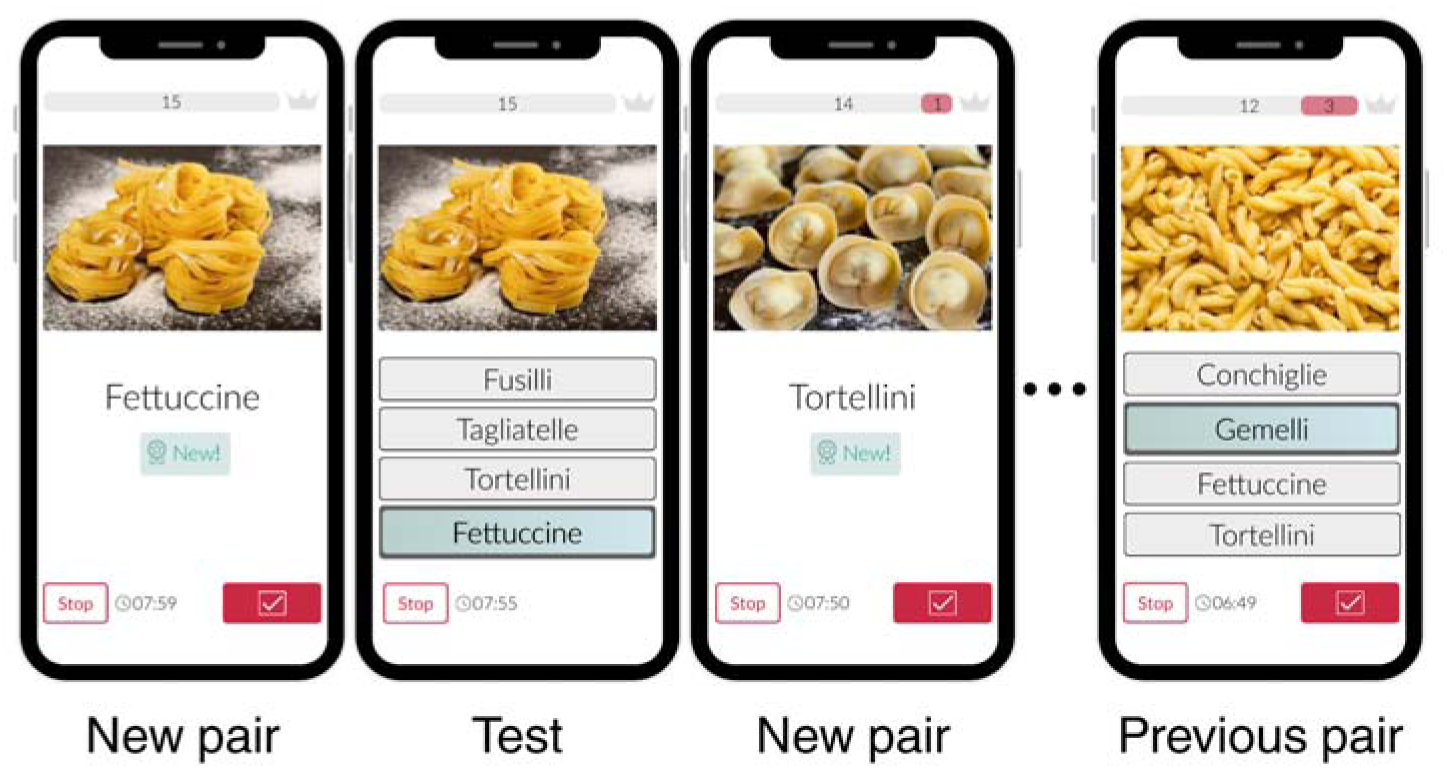
Interface of the memory assessment software. Examples of the memory task in which participants engaged in adaptive retrieval practice, alternating between memorizing new pairs of items and being tested on previously encoded pairs.

### Episodic Memory Model

The model used herein was originally developed by Anderson & Schooler (*16*) and later expanded by Pavlik and Anderson (*17*) and van Rijn (*18*). Consistent with the Multiple Trace Theory (*19*), the model assumes that all memories are created by the accumulation of individual episodic traces encoded every time the same information is encountered (Fig 1A). Each trace independently decays over time according to a power law (*29*). The odds of retrieving a memory *m* at time *t* are proportional to its activation *A*(*m*, *t*), which represents the log odds of remembering any of its component traces, as shown in Eq. (1).

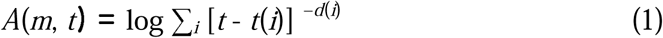

Where *t*(i) is the creation time of the *i*-th trace and *d*(*i*) is its characteristic power decay rate. This trace-specific decay rate, in turn, depends on the residual activation of the memory at the time the trace was created (*17*, *22*), plus a fixed component φ:

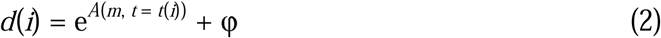

where φ is the *Speed of Forgetting* (SoF). Because Eq. (2) makes the decay rate of each trace dependent on the memory’s activation, it provides an explanation for the spacing effect (*17*): traces closer in time have higher decay rates because of the memory’s greater activation *A*(*m*, *t*) at time *t*(*i*). Thus, in this model, the odds of being able to recall a memory at a later time depend solely on the rate at which the memory is forgotten. Additionally, this suggests that by looking at the history of a memory and the number of times it has been assessed, it is possible to determine the rate at which that memory is forgotten.

The adaptive learning system exploits the fact that, in the model, the activation of a memory *A*(*m*, *t*) translates directly to its probability of a correct answer to a test trial as well as the associated response time. Specifically, since *A*(*m*, *t*) represents the log odds of retrieving a memory, the probability of a correct answer *P* is expressed as:

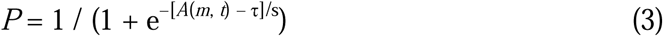

where τ is the activation threshold beyond which a memory cannot be retrieved any longer, and *s* is activation noise. The expected response time *RT* to retrieve a memory is given by:

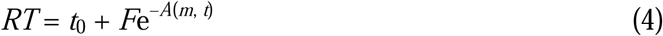

where *t*_0_ is a free parameter that represents the temporal costs associated with non-memory related processes, such as motor preparation and perceptual encoding, and *F* is an individual parameter that reflects speed of processing.

### Study Materials

Fifty-three lessons were prepared covering a wide range of topics (e.g., European capitals, Swahili words, Asian flags, bird species, types of pasta, and flower species). Topics were selected to appeal to a broad audience and encourage ongoing participation, with an emphasis on variety and general interest. A key criterion for inclusion was that each topic could support the creation of at least 15 distinct item pairs. For example, the “Pasta” lesson was included because there are more than 15 recognizable types of pasta that could be meaningfully paired with their names. While the content selection process was not strictly systematic, it was intentionally diverse and guided by the expectation that topics with broad, accessible appeal would be most engaging for repeated weekly participation. All lessons were internally tested prior to deployment: the lead author and research assistants completed each lesson to ensure that the material was appropriately calibrated (neither too difficult nor too easy) for the 55 to 85 year-old population. Each lesson contained 15 object–label pairs; objects were presented as images (e.g., a photo of “Fettuccine” in the Pasta lesson) in half of the lessons and as words (e.g., “France” paired with “Paris” in the European Capitals lesson) in the other half. Because of the adaptive nature of the paradigm, the number of items studied during each session varied across participants, depending on their response times and error rates: participants with lower memory function would naturally see fewer items that are repeated a larger number of times. Independently of the number of items learned and their repetitions, all sessions had a fixed duration of 8 minutes. Pilot work has demonstrated that this duration finds a balance between minimizing the time demands on our participants while still allowing the model parameters to stabilize, with the difference between groups remaining stable even as the parameters for newly introduced facts are being estimated (see Supplementary Fig 1).

Of the 53 lessons, 49 used a four-option multiple-choice format, while 4 employed verbal recall. All verbal recall lessons were image-based. In these sessions, conducted via videoconferencing, participants were shown a visual cue and asked to provide the associated response aloud.

### Procedures

Participants were invited for weekly sessions until drop out. Each week’s lesson was the same for all participants but, since participants enrolled on a rolling basis, each of them saw a different sequence of lessons. If a participant did not complete a lesson during the allocated week, they were given the opportunity to complete the lesson after they completed the full cycle of 52 or 53 weeks.

In addition to these multiple-choice sessions, participants were invited, at approximately three-months intervals, to participate in four verbal recall sessions. In these sessions, the cue was still presented on the screen but participants were asked to answer verbally. All recall tests were conducted over a videoconferencing platform, with verbal responses automatically transcribed through Google’s speech-to-text software (*30*). Only one participant opted out of the recall-based sessions. On average, the remaining 50 participants contributed 3.1 recall-based performance measures each.

### Data Processing

The activation and SoF values for each item were calculated using the computational cognitive memory model. The mean SoF values for each user were identified by using the last SoF value of each item at the very last repetition of that item. Interpretable SGMA scores were then generated by passing mean SoF values through a sigmoid function whose inflection point corresponded to 100. The data was then filtered to only contain the first full session of a lesson (> 6 min). This was needed to eliminate any superfluous sessions (some participants desired to complete the task more than once). The data was also organized by the week the lesson was completed to view temporal trends. All of the analyses were performed in R (v4.2.2); linear mixed-effect models were implemented with the *lme4* package (*31*).

## Results

### Model Goodness-of-Fit

Before analyzing the results of a model-based memory assessment, it is important to examine how well the model itself captures individual differences in participants’ accuracies and response times (*32*). To do so, we computed the predicted activation for each trial of every lesson from every participant by entering each participant’s history of previous presentations and the final participant-specific φ per fact into Equations 1 and 2.

Trial-by-trial predicted accuracy was derived from the estimated memory activation using Equation 3. The parameters *s* and τ were estimated separately for each participant and lesson via a logistic regression model of the form *y* = 1 / (1 + e^−β0−β1*x*^). Here, *y* represents an observed trial accuracy, and *x* the corresponding memory activation *A*(*m, t*). From Equation 3, it follows that −β_0_ − β_1_*x* = −*A*(*m, t*) / *s* + τ / s and, therefore, τ = β_0_ / β_1_ and *s* = 1 / β_1_.

Due to the assessment’s adaptive nature, participants answered most trials correctly (mean accuracy: 89%; range: 57.4%–98.7%). Because of this, our evaluation of model fit focused on the model’s ability to predict each participant’s *mean* accuracy across lessons rather than single-trial classifications, which would be subject to ceiling effects. Fig 4A plots each participant’s predicted mean accuracy against their observed mean accuracy. Observed and predicted accuracies were significantly correlated across both MCI patients (*r*(24) = 0.72, *p* < 0.001) and healthy controls (*r*(27) = 0.56, *p* < 0.01).

**Fig 4:**
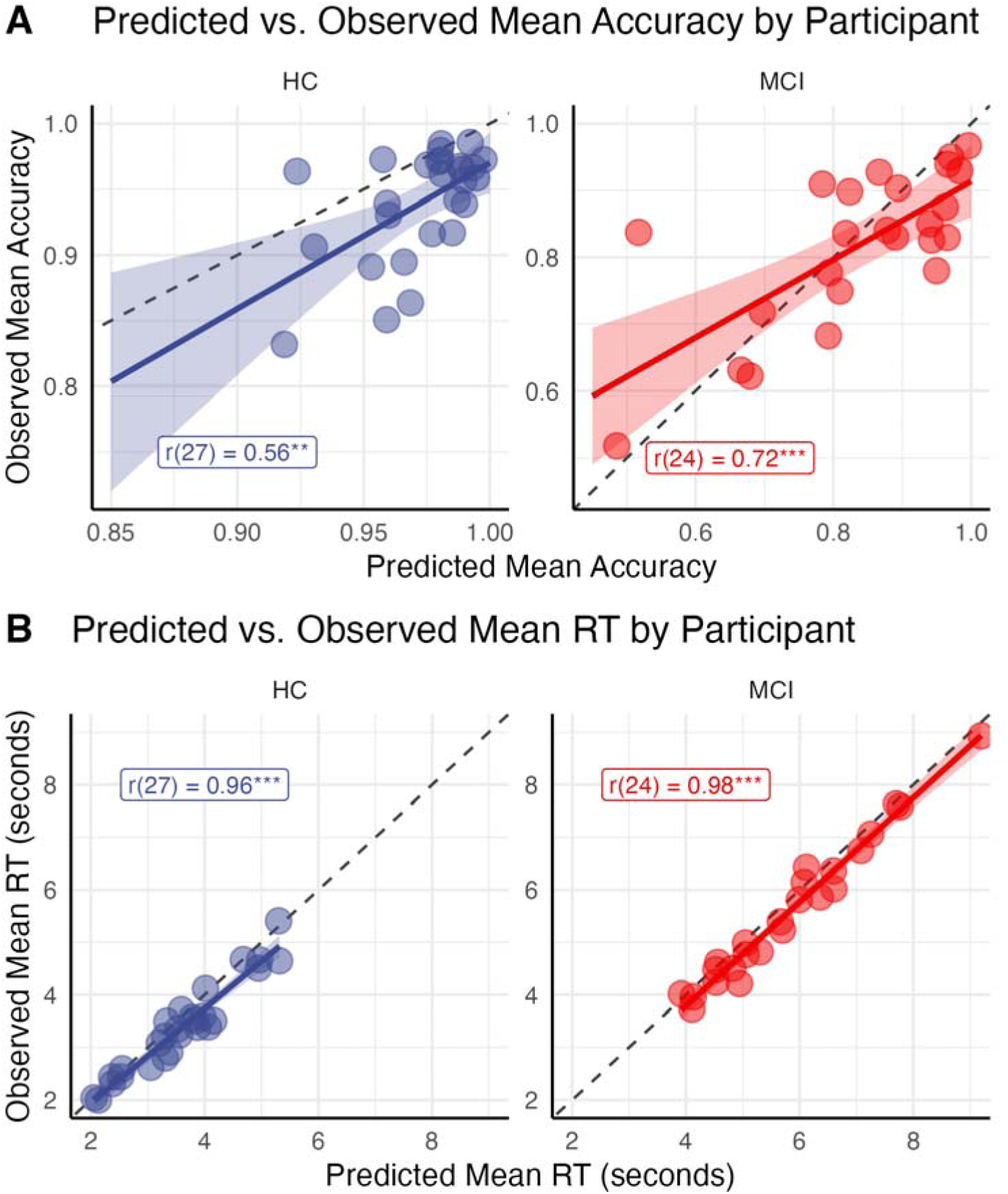
Model fit for individual participants. (A) Mean participant accuracy of each participant plotted against the corresponding accuracy predicted by the model. (B) Mean response time for each participant plotted against the corresponding model prediction. Model predictions for accuracy and RT are based on the individual mean speed of forgetting estimated in the task. *** *p* < 0.001, ** *p* < 0.01, * *p* < 0.05.

Fig 4B similarly plots the observed response times for each participant against the corresponding model predictions, computed by averaging the trial-level predictions using Equation 4. Again, the model’s predictions closely match the observed data for both MCI patients (*r*(24) = 0.98, *p* < 0.001) and healthy controls (*r*(27) = 0.96, *p* < 0.001). Unlike accuracies, however, response times are continuous and can also be meaningfully investigated at the trial level. To do so, we first excluded 2.5% of trials with response times below 300 ms or above 15 s, which likely reflect accidental responses or momentary disengagement from the task, leaving a total of 174,223 trials.

Another common way to evaluate a model’s fit is to compare the model’s predicted response times distribution against the observed distribution of response time for each participant (*33*). The similarity between the two distributions can be visually assessed through a decile plot, that is, by plotting the predicted RT in each decile against its empirical counterpart. Fig 5 depicts the predicted vs. observed values of the first nine deciles of each participant’s RT distributions; for visual clarity, the last decile is omitted, as it often includes spurious response outliers that the model cannot reproduce (for completeness, the full decile plot is provided in Supplementary Fig 2). As the figure shows, almost all decile points tend to fall along the identity lines—a sign of the similarity of the distributions.

**Fig 5.**
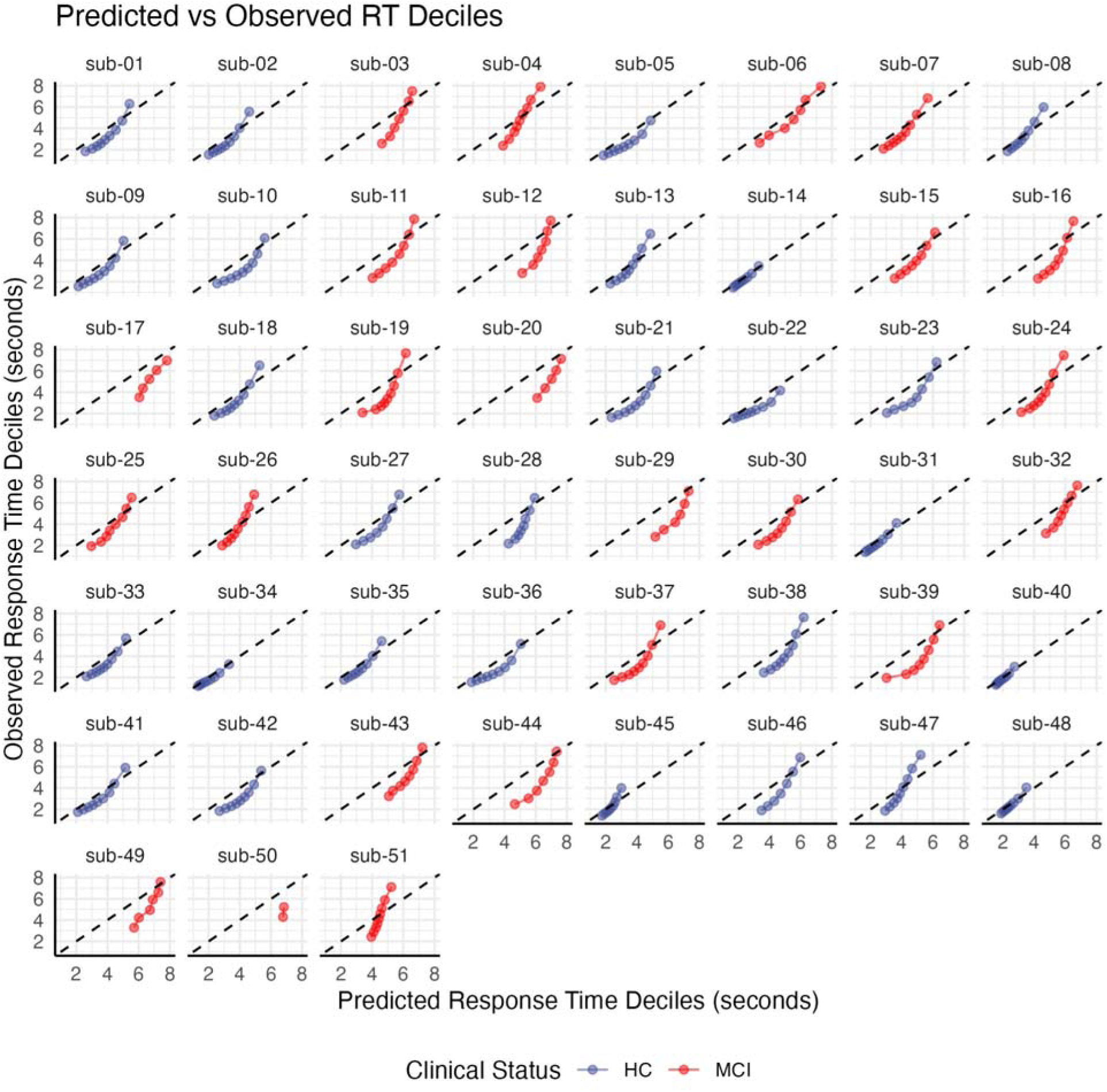
Model fit for individual response times. Decile plots, showing the predicted vs. observed RTs corresponding to the response in the 1^st^, 2^nd^, … 9^th^ decile for each participant.

The similarity between observed and predicted response time distributions can also be assessed using Kullback–Leibler (KL) divergence and χ^2^ tests. KL divergence reflects the average information loss when predictions replace observed data, while χ^2^ tests assess the goodness of fit to the observed frequency distribution; in both cases, lower values indicate better fit. To compute both metrics, response times were binned in 1-second intervals and predicted vs. observed counts were compared. Both statistics, together with the predicted and observed distributions for all participants, are depicted in Supplementary Fig 3. The mean KL divergence was 1.37 across participants and 0.30 across all trials. The mean χ^2^ value was 131.32, with *p* > 0.05 for all but one participant. For all trials combined, the χ² test also indicated a good fit (χ²(196) = 132, *p* > 0.23). Together, these analyses support the model’s validity in predicting both accuracy and response times across individuals.

### Stability and Reliability of Model-Based Assessment Scores

Having established that the model adequately captures the individual behavior of our participants, we can now examine the validity of the model-derived SGMA scores.

Participants completed a total of 1,968 recognition tests, with an average of 38.6 weekly assessments per individual (range 11–46). Although they were allowed to leave the study at any time, participants remained engaged for an average of 48.5 weeks (range 22–53); no differences were found between the mean engagement of controls (51.0 ± 4.0 weeks) and that of MCI patients (48.0 ± 8.2 weeks; *t*(32.31) = 1.66, *p* = 0.11). Despite using vastly different lesson topics, the mean weekly SGMA scores exhibited minimal variability (Mean = 75.2 ± 5.0; range 67.51 – 88.38). Across all pairs of lessons, SGMA scores exhibited a mean Pearson pairwise correlation coefficient of *r* = 0.70, indicating consistent inter-lesson reliability (Fig 6A-B). Because this effect could be partially driven by the stable difference between groups, we also replicated the analysis separately for patients and controls. These separate analyses found slightly lower, but still reliable inter-lesson correlations for MCIs (*r* = 0.56) and controls (*r* = 0.65), in line with other findings (*34*).

**Fig 6.**
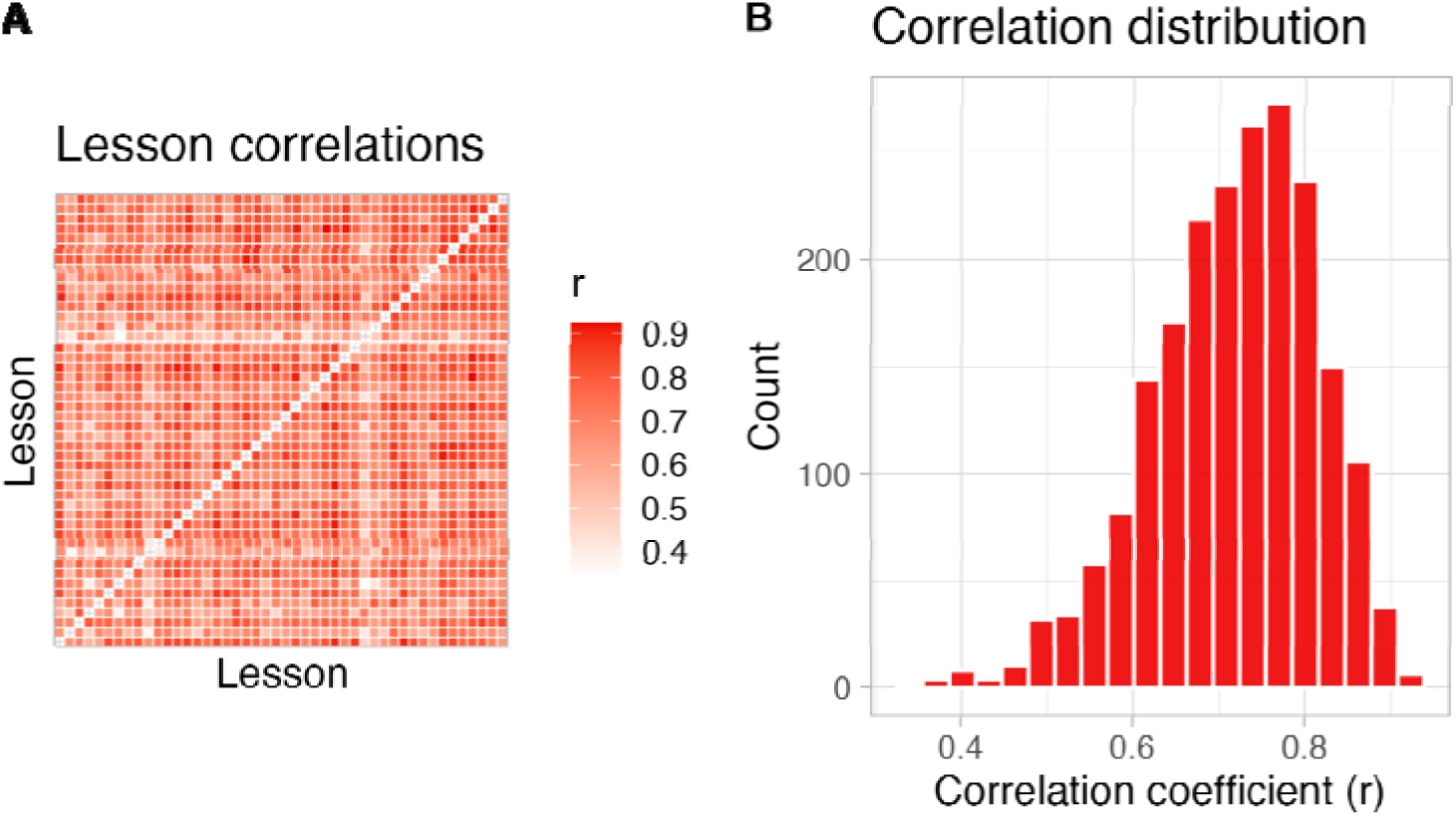
Reliability and stability of SGMA score. **(A)** Heatmap of Pearson pairwise correlations (*r*) between participants’ SGMA scores across different lessons. **(B)** Distribution of the pair-wise correlation coefficients from panel A.

### Longitudinal Group Differences in Model-Based Assessment Scores

As expected, SGMA scores were lower in the MCI group (63.6 ± 9.4) than in controls (83.3 ± 13.2; Fig 7A-B). This raw difference, however, does not account for potential confounds, such as differential group effects of practice or individual differences. To separate out these confounds, individual SGMA scores from all lessons were analyzed using a linear mixed-effects model. The model included fixed effects for participant group (MCI vs. controls; dummy-coded: MCI = 1, Controls = 0), the temporal order of each lesson within a participant’s schedule (weeks since enrollment), and their interaction. Sex (contrast-coded; male = 0.5, female = –0.5), age (centered), and stimulus format (text vs. image, contrast-coded) were included as covariates, along with a group-by-age interaction. To account for within-subject and within-lesson variability, the model included random intercepts and random slopes for week for each participant, as well as random intercepts for each lesson. This structure allowed us to model individual learning trajectories (i.e., participant-specific slopes) while adjusting for differences in baseline SGMA performance and lesson difficulty. Table 1 provides a full account of the model’s results. After controlling for all covariates and random effects, a significant (*p* < 0.001) and large (12.36 points) difference between groups remained. The temporal order of lessons also showed a significant (*p* < 0.001) positive fixed effect, reflecting a modest average practice effect of 0.17 points per week, or a 0.2% increase per assessment. Critically, this practice effect interacted significantly with group (*p* < 0.001), indicating that the average slope of SGMA improvement over time was reliably smaller (by 0.10 points per week) in the MCI group compared to controls (Fig 7C). Additionally, age and sex also had significant effects, with SGMA scores being 7.61 points lower in males (*p* = 0.019) and declining by 0.91 points per year (*p* = 0.001). The specific nature of the stimulus format, on the other hand, had no discernible effect on memory performance (*p* = 0.55).

**Fig 7.**
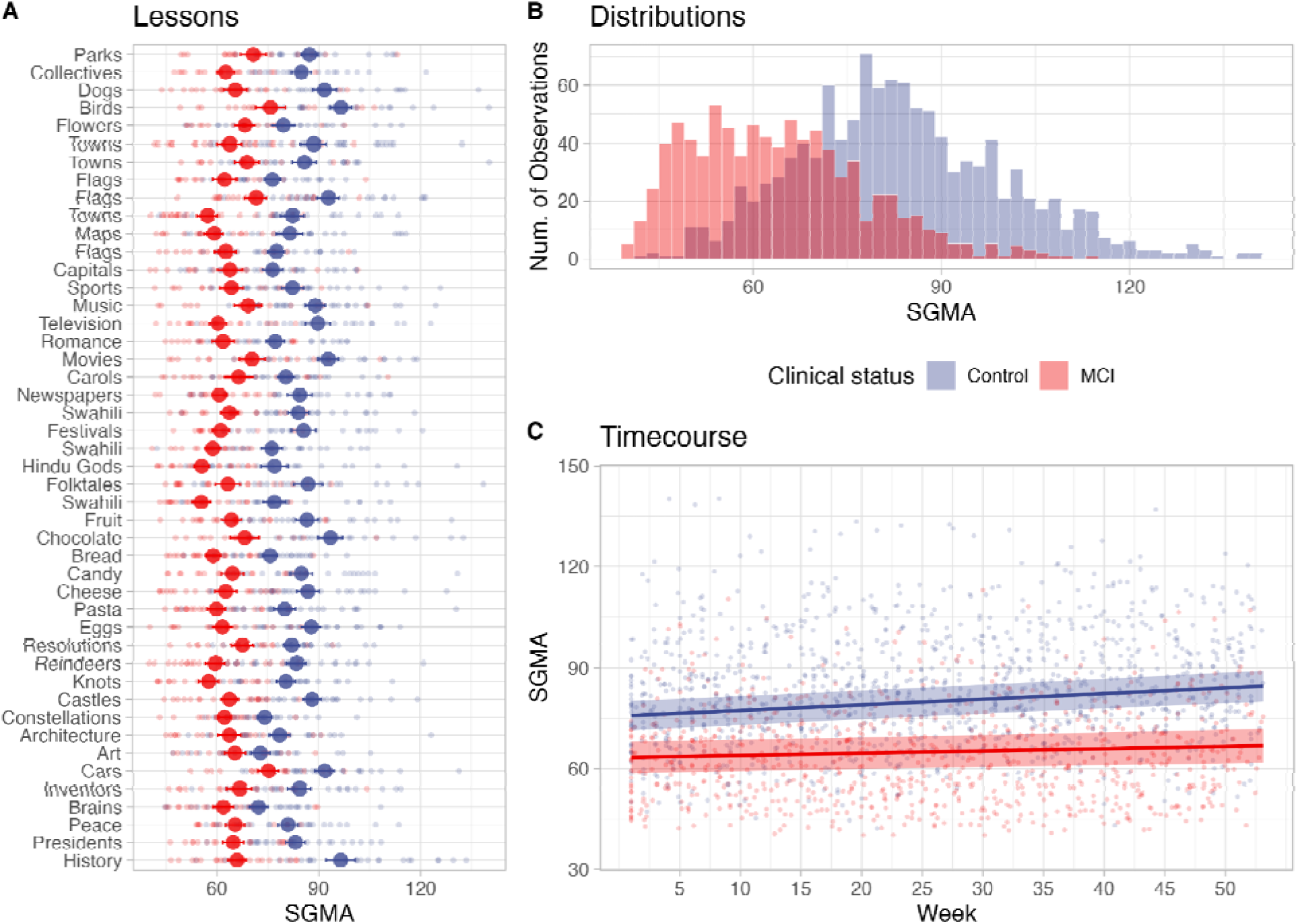
SGMA scores across lessons, groups, and over the timecourse of the study. **(A)** Individual SGMA scores per lesson, color-coded by group (Control: blue, MCI: red). **(B)** Density plot of SGMA scores. **(C)** SGMA scores over time, with points representing individual weekly scores. Trend lines show estimated group-level trajectories derived from a linear mixed-effect model that includes random intercepts for lessons and users, and random slopes for week by user. Shaded bands represent 95% confidence intervals around the predicted fixed effects.

**Table 1:** Results of the linear mixed-effect model analysis of the recognition memory assessments.

|  |  | **SGMA Score** |  |
| --- | --- | --- | --- |
| *Predictor* | *Estimate* | *95% CI* | *p* |
| (Intercept) | 79.45 | 74.78 – 84.12 | < 0.001 |
| Group (Dummy coded) | -12.36 | -18.81 – -5.91 | < 0.001 |
| Week | 0.17 | 0.12 – 0.22 | < 0.001 |
| Age in Years (Centered) | -0.91 | -1.44 – -0.39 | 0.001 |
| Sex (Contrast coded) | -7.61 | -13.98 – -1.24 | 0.019 |
| Format (Contrast coded) | 0.88 | -2.06 – 3.82 | 0.557 |
| Group × Week | -0.10 | -0.17 – -0.03 | 0.004 |
| Group × Age | 1.68 | 0.71 – 2.66 | 0.001 |
| **Random Effects** |  |  |  |
| $\sigma^2$ | 86.46 | | |
| Participant (Intercept) | 92.63 |  |  |
| Lesson (Intercept) | 23.26 |  |  |
| Participant (Slope) | 0.00 |  |  |
| ICC | 0.24 |  |  |
| $N_{\text{Lesson}}$ | 46 | | |
| $N_{\text{Participant}}$ | 51 | | |
| *Observations* | *1,968* |  |  |
| *Marginal R<sup>2</sup>* | *0.556* |  |  |
| $Conditional R^2$ | 0.662 | | |

### Diagnostic Accuracy of Model-Based Assessment Scores

Next, we investigated whether model-based SGMA scores are diagnostically informative. To address this question, individual SGMA scores from all sessions were divided into bins ranging from 30 to 150 in increments of one; for each bin, we computed the proportion of scores coming from MCI individuals. The probability of an MCI diagnosis reliably declined as a logistic function of the SGMA score, with an inflection point at the value of 69 (Fig 8A). This trend was well-fitted by a simple logistic model, which accounted for 38.8% of the variance (Nagelkerke’s R²).

**Fig 8.**
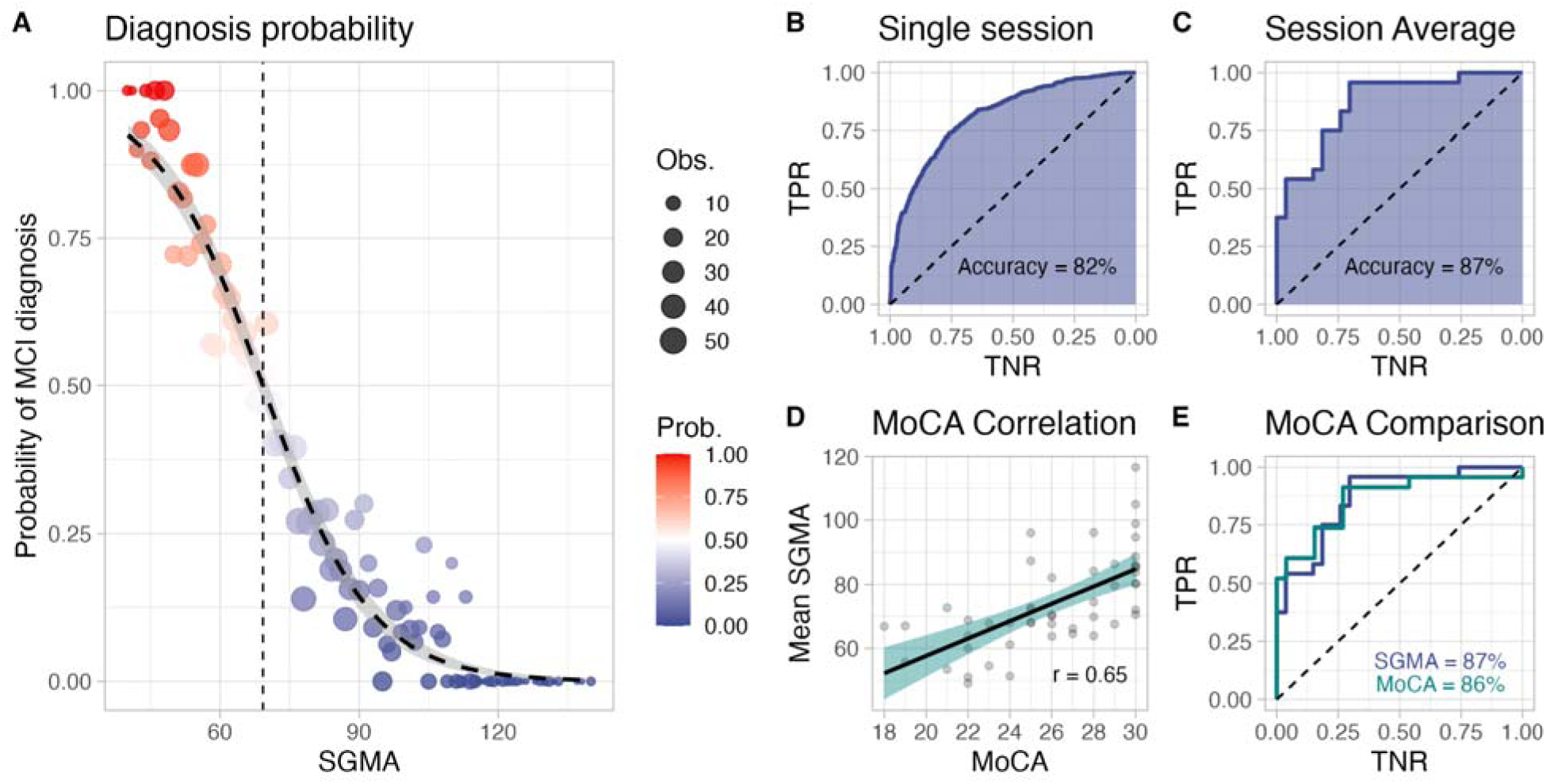
Diagnostic properties of computational phenotyping. **(A)** Probability of an MCI diagnosis for an observed SGMA score, with point size indicating the number of observations associated with that SGMA score. The thick dashed line represents a logistic regression predicting the probability of an MCI diagnosis based on SGMA score, with the shaded area representing the 95% confidence interval around the fitted logistic regression curve. Red color represents higher probabilities of MCI. The vertical line marks the SGMA threshold where MCI probability reaches 50%. **(B)** ROC curve evaluating the SGMA’s ability to discriminate between MCI and controls from a single session (out-of-sample AUC = 82%). **(C)** ROC curve for the average SGMA index per participant across multiple sessions, showing improved discrimination accuracy (out-of-sample AUC = 87%). **(D)** Scatter plot of SGMA scores against MoCA (Montreal Cognitive Assessment) scores. **(E)** ROC curves comparing the discrimination accuracy of SGMA and MoCA scores, showing similar performance (SGMA = 87%, MoCA = 86%). The dashed lines in B, C, and E represent the baseline of random predictions, TNR is True Negative Rate, TPR is True Positive Rate.

To further examine the discriminatory power of our model-based approach, we trained a logistic classifier to predict an individual’s diagnosis (MCI or control) using the SGMA score as its sole input. The classifier was applied to all of the 1,968 session scores and cross-validated using a leave-one-out scheme. Its out-of-sample classification performance, measured as the area under the curve (AUC) of its Receiver Operator Characteristic (ROC), was 0.82 (Fig 8B). In other words, a single 8-min sessiondiscriminated MCI from Control cases with an out of sample AUC of 0.82. We also applied the same model and cross-validation procedure to a reduced dataset that contained only the mean SGMA scores per participant. The average out-of-sample discrimination accuracy on the unseen participant was 87% (Fig 8C).

To put these numbers into perspective, the performance of the single-predictor SGMA classifier was compared against an analogous classifier based on the results of the Montreal Cognitive Assessment (MoCA), the most commonly used, physician-administered screening test for MCI diagnosis (Fig 8D; (*35*)). In our sample, MoCA and SGMA scores exhibit a correlation of *r* = 0.65 (*p* < 0.001). The out-of-sample diagnostic accuracy of MoCA was 86%, not significantly different from the average diagnostic accuracy of SGMA. Remarkably, although the MoCA test assesses a wider range of cognitive domains than SGMA, it does not outperform it in classification accuracy (Fig 8E). This suggests that SGMA compensates for the reduced set of abilities tested by achieving greater precision in assessing memory function.

Because sex was a significant factor affecting SGMA scores in our statistical analysis, we examined whether its inclusion in the logistic classifier would significantly alter SGMA model-based assessment. An extended (SGMA + Sex) classifier did not perform significantly better when classifying participants either using single sessions (0.87 vs. 0.82) or the session average (0.85 vs. 0.87). Using the same logic, we also examined whether adding mean response times or mean accuracies would affect classification.

### Relationship Between Recall and Recognition Assessments

One of the drawbacks of typical assessments is that, being based entirely on test accuracy, they cannot distinguish between deficits in *accessing* a memory (due, for example, to frontal lobe damage and executive dysfunction) and true memory loss. In contrast, SGMA is based on a model that explicitly captures forgetting and memory loss; thus, SGMA is potentially immune to this confound. A comparison of recall vs. recognition tests offers the opportunity to empirically test this claim (Fig 9A-D). Because they pose additional demands on executive function, recall tests are associated with lower accuracy. However, they also lead to better processing and longer retention (*36*). Therefore, we expect SGMA to yield higher scores in memory recall than in memory recognition paradigms, despite a predictable drop in accuracy.

**Fig 9.**
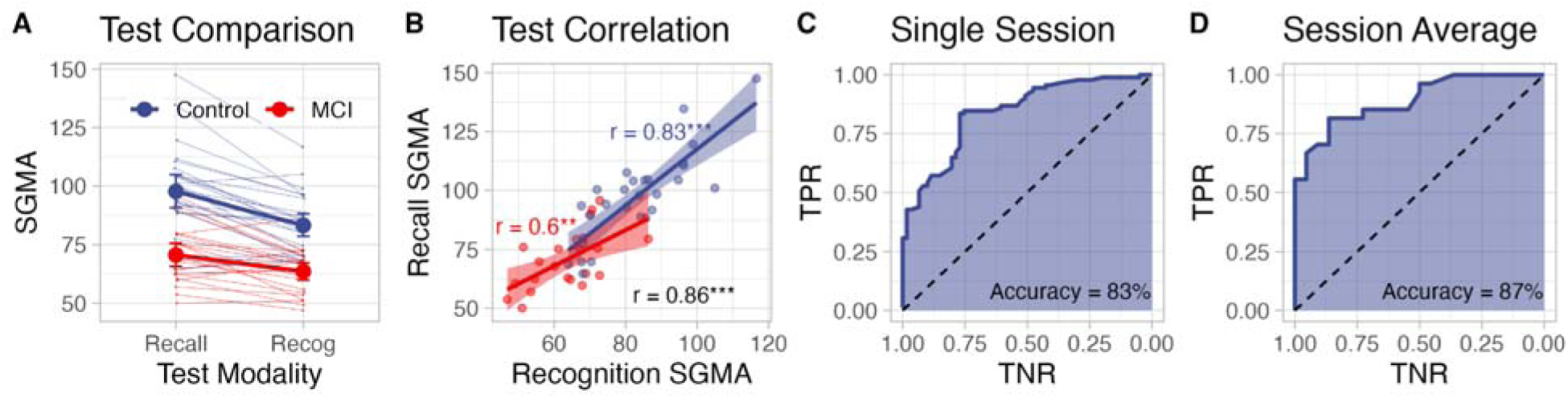
Relationship between recall and recognition memory performance. **(A)** SGMA scores for recall and recognition tests, with lines connecting individual participants. Controls shown in blue, and MCI in red. **(B)** Scatter plot showing the relationship between recognition and recall SGMA scores, with linear regression lines for each group. Shaded areas represent 95% confidence intervals. **(C)** ROC curve evaluating the discrimination accuracy of a single SGMA measurement (out-of-sample AUC = 83%). **(D)** ROC curve showing improved discrimination accuracy when using the average SGMA index across multiple sessions (out-of-sample AUC = 87%). TNR = True Negative Rate; TPR = True Positive Rate; \*\*\**p* < 0.001, ** *p* < 0.01, * *p* < 0.05

To compare recall vs. recognition scores and control for confounding variables, we computed the mean recognition and recall scores for each participant and entered them into a linear mixed-effectmodel. The model included the fixed effects of test modality (recall vs. recognition), the participant group (MCI vs. control) and their interaction. As in the recognition analysis, sex and age were included as confounding variables together with a group-by-age interaction. Finally, the model included random intercepts for each participant to account for individual differences in baseline performance and a random slope to account for differences in sensitivity to test modalities. The resulting model, described in Table 2, accounted for 85% of the variance. After accounting for all confounds and interactions, the model identified a significant (*p* < 0.001) group difference, with the MCI group scores being 15.3 points lower than controls on average (i.e., across recall and recognition; Fig 9A).

**Table 2:** Linear mixed-effect model analysis of individual mean SGMA scores across testing modality.

|  |  | **SGMA Score** |  |
| --- | --- | --- | --- |
| *Predictor* | *Estimate* | *95% CI* | *p* |
| (Intercept) | 76.87 | 68.01 – 81.73 | < 0.001 |
| Group (Dummy coded) | -15.33 | -23.06 – -7.59 | < 0.001 |
| Test Modality (Dummy coded) | 14.36 | 10.36 – 18.36 | < 0.001 |
| Age (Mean centered) | -0.93 | -1.52 – -0.34 | 0.002 |
| Sex (Contrast coded) | 9.77 | 2.57 – 16.97 | 0.008 |
| Group × Modality | -7.11 | -13.06 – -1.16 | 0.020 |
| Group × Age | 1.97 | 0.87 – 3.07 | 0.001 |
| **Random Effects** |  |  |  |
| $\sigma^2$ | 54.85 | | |
| Participant (Intercept) | 92.21 |  |  |
| ICC | 0.24 |  |  |
| $N_{\text{Participant}}$ | 51 | | |
| *Observations* | 100 |  |  |
| *Marginal R<sup>2</sup>* | 0.606 |  |  |
| *Conditional R<sup>2</sup>* | 0.853 |  |  |

Importantly, recall scores were significantly *higher* (by 14.3 points, *p* < 0.001) than recognition scores—despite the mean recall accuracy being *lower* than recognition accuracy (67.2% vs. 90.4%). This result, therefore, confirms our hypothesis that our model-based SGMA scores indeed capture the underlying dynamics of consolidation, rather than confounding consolidation and retrieval failures. This is further confirmed by an analysis of the within-session variability across the different items studied. If the memory problems of the MCI patients were due to an increase in the frequency of retrieval failures for certain items, rather than an accelerated forgetting across the board, we would expect to find greater within-session item-level variability in patients than in controls. Instead, we find the exact opposite: within-session variability, operationalized as the standard deviation of SGMA scores across all items, was higher in healthy controls (*M*= 29.9, *SD* = 3.0) than in MCIs [*M* = 22.4, *SD* = 6, *t*(32.96) = 5.55, *p* < 0.001]. Finally, the fact that lower SGMA scores reflect impaired consolidation rather than impaired retrieval is supported by the lack of difference in SGMA scores between different subpopulations of MCI patients. If SGMA scores were confounded by retrieval deficits, we would expect amnestic patients with deficits in multiple domains to score lower than patients with exclusive long-term memory impairments, since concurrent visual, language, or executive function problems also impair retrieval process. Instead, we found no difference between the two groups [*M* = 65.57, *SD* = 8.97 vs. *M* = 59.60, *SD* = 9.53, respectively: *t*(14.24) = 1.42, *p* = 0.18].

In addition, a significant (*p* < 0.001) group by modality interaction showed that, compared to controls, the MCI group had a smaller (7.11 points less) benefit from the recall modality; this interaction also suggests that the model is sensitive to the MCI group’s difficulties in encoding and consolidating information and, therefore their limited benefit from deeper processing in recall paradigms. Finally, age and sex were also found to play a significant role, with SGMA scores being 9.77 points lower in males (*p* < 0.01) and declining by 1.97 points per year (*p* < 0.001).

Despite theoretical distinctions between recall and recognition memory (*37*), SGMA scores exhibited a strong correlation across all participants for both test types (Pearson *r* = 0.86, *p* < 0.0001; Fig 9B). This further confirms that SGMA scores capture a common underlying process of memory consolidation that is common to both modalities.

Finally, an analysis of the recall sessions’s diagnostic sensitivity revealed comparable classification accuracy to recognition-based SGMA, as indicated by out-of-sample ROC AUC values of 83% for a single-session (Fig 9C), and 87% for the mean (Fig 9D).

### Relationship to Other Neuropsychological Assessments

Lastly, we examined the relationship between SGMA scores and other common neurocognitive assessments. All participants underwent extensive neuropsychological assessments at the local Alzheimer’s Disease Research Center. The assessment included all of the tests in the NACC Uniform Data Set (UDS) Version 3 (*38*): the MoCA; the Craft Story 21, immediate and delayed (*39*), the Benson Complex Figure Test, immediate and delayed (*40*); the forward and backward number span; the letter list generation task, using F and L letters; the category list generation task, using Animals and Vegetables categories; the Trail Making Test, parts A and B (*41*); the Multilingual Naming Test (MINT: (*42*)); and, finally, the memory score and the sum-of-boxes scores from the Clinical Dementia Rating (CDR: (*43*)).

Fig 10 provides an overview of the Pearson correlations of these neuropsychological tests with SGMA scores, while Supplementary Fig 4 provides the full correlation matrix between all tests. In line with their diagnostic accuracy, SGMA scores were robustly and inversely associated with both the memory (*r* = –0.56, *p* < 0.001) and the sum-of-boxes scores (*r* = –0.71, *p* < 0.001) of the CDR. As expected, SGMA scores were significantly and positively correlated with the outcomes of tests that also tap into long-term memory function, such as the immediate (*r* = 0.51, *p* < 0.001) and delayed recall (*r* = 0.50, *p* < 0.001) of the Craft Story test, the delayed recall of the Benson complex figure (*r* = 0.34, *p* = 0.03), the letter list generation tasks (letter F: *r* = 0.46, *p* = 0.002; letter L: *r* = 0.47, *p* = 0.002; Both F and L: *r* = 0.49, *p* = 0.001), the category list generation tasks (vegetable names: *r* = 0.57, *p* < 0.001), as well as with tests of verbal working memory (backwards number span: *r* = 0.39, *p* = 0.01). Although only marginally significant, the correlations with the list generation test for the animal category (*r* = 0.241, *p* = 0.1) and with the forward number span (*r* = 0.26, *p* = 0.09) were similarly positive.

**Fig 10.**
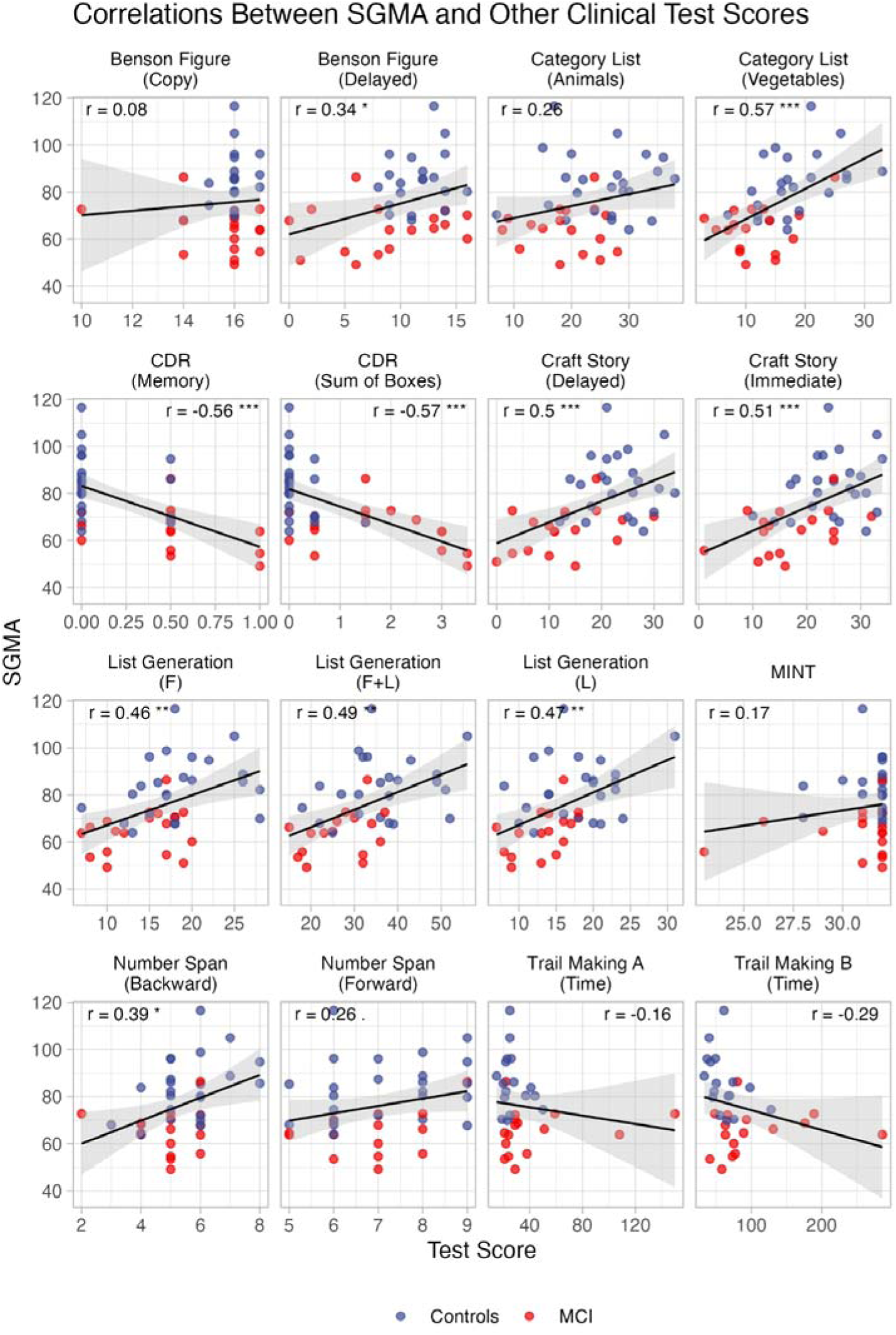
Correlations between SGMA and other clinical assessments. Panels represent scatterplots between individual SGMA scores and other test scores; lines and shaded areas represent regression lines and standard errors of the estimate. \*\*\**p* < 0.001, ** *p* < 0.01, * *p* < 0.05;. *p* < 0.1

Conversely, SGMA scores were *not* significantly correlated with tests that tap into distinct cognitive abilities, such as simply copying the Benson complex figure, which requires visuo-spatial skills but no use of long-term memory (*r* = 0.08, *p* > 0.64); the Trail Making Test, which assesses set-shifting and executive functions (versions A: *r* = –0.16, *p* = 0.38; version B: *r* = –0.28, *p* = 0.11); and the MINT, which assesses linguistic abilities (*r* = 0.17, *p* = 0.30).

## Discussion

Taken together, the results confirm that the Seattle-Groningen Memory Assessment (SGMA), a new test based on computational phenotyping, can successfully monitor memory function over a period of one year in a cohort of representative elderly individuals with either intact or mildly impaired cognitive function. Lower SGMA scores were reliably and proportionally associated with greater probabilities of MCI diagnosis, so that our 8-minute, online assessment could achieve an out-of-sample diagnostic accuracy of 87%, an accuracy that sits at the maximum of the range identified in earlier work (*44*, *45*) and is comparable to that of widely used traditional tests, such as the Montreal Cognitive Assessment. In addition, our model-based SGMA score was found to be repeatable and stable across weekly sessions, materials, and even testing modalities, confirming the validity of the underlying model and its ability to capture a latent aspect of memory function that generalizes across task formats.

### Relationship to Other Digital Assessment Tools

SGMA is not the only digital assessment tool that has been proposed to detect memory impairments. In fact, the pressure to scale assessment at a population level and to detect memory impairments early has led to a proliferation of digital tools (see (*46*) for a recent review). A number of solutions exist, including digital ports of well-established paper-and-pencil instruments (such as digital versions of the Montreal Cognitive Assessment (*47*) or the CANTAB suite (*48*)) and entirely novel paradigms, based a suite of task that reflects more contemporary views of cognition long-term memory function (e.g., Cognitron (*49*), CogState (*50*, *51*), Neotiv (*52*), or Neurotrack (*53, 54*); see also (*55*), for a comparison to the Amsterdam Cognition Scan digital batteries).

Compared to these tools, our approach achieves similar specificity and sensitivity, but allows greater simplicity and interpretability, being based on a simple, intuitive, and well-established paradigm to investigate memory (i.e., paired associates). Critically, SGMA shows greater test-retest reliability than any of the other products, and allows much more frequent testing. Most importantly, our tool is unique because of the nature of its assessment. To the best of our knowledge, it is the only assessment tool based on computational phenotyping and a “digital twin” approach. Unlike other tools, SGMA goes beyond simple accuracies and reports interpretable parameters. The SGMA score is therefore relatively independent from the stimuli, difficulty, and exposure history that produced them. This facilitates longitudinal use, since the raw scores reported by other tools are difficult to disentangle from item difficulty and from the familiarity and sensorimotor gains that accompany repeated testing. As our results show, our approach affords greater flexibility of use, including the possibility of quickly adapting the materials to new languages or even designing new culturally-appropriate stimuli on the fly.

A recent development in the field has been the emergence of AI-based tools for quick and accurate diagnosis (e.g., (*56–58*). These tools, often based on small samples of speech recordings (*58*), are particularly attractive because of their ease of use and unobtrusiveness. One of the limitations of such models is their restricted diagnostic scope: they are explicitly designed as classifiers for specific conditions (e.g., Alzheimer’s dementia). In contrast, SGMA is fundamentally trans-diagnostic: it is designed first and foremost to reliably assess long-term memory function, and, although this feature makes it potentially diagnostic for certain conditions (including Alzheimer’s), it can be applied widely and provide important information across a variety of pathologies. Another limitation of AI-based approach is their limited explainability: it is often impossible to precisely understand which features the underlying model has latched on to inform its decision. SGMA, on the other hand, is based on an interpretable cognitive model whose parameters are intrinsically explainable. Finally, the relative simplicity of SGMA’s underlying model (which contains only the five parameters of Equations 3 and 4, instead of the million or billion parameters underlying most AI-based models) allows reliable estimates from very limited data and without any underlying training dataset.

The latter point underscores another unique characteristic of SGMA, its natural security and privacy. Not only does SGMA not require storing massive amounts of patient data for training purposes, but, because it is adaptive and the model’s central parameter is re-estimated after every response, it does not require maintaining *any* persistent behavioral data from the participant. This allows much greater security and protection of patient privacy and confidentiality and might facilitate its adoption in critical domains as it meets stringent AI regulations.

### Relationship to Other Model Parameters

Although the model-fitting procedure was carried out with the sole intent to examine the model’s ability to reproduce the behavior of our participants, it might also provide important clues about the nature of the cognitive differences between patients and controls. Indeed, we found important differences between patients and controls in the parameters that govern how memory activation translates into response times, that is, the non-retrieval time *t*_0_ and the latency factor *F* (Supplementary Fig 5). It is important to stress that these ancillary parameters were estimated across all sessions, and only *after* the determination of the speed of forgetting; as such, they might be potentially confounded with our SGMA score and lack its temporal resolution (see (*59*) for an alternative approach). Nonetheless, and despite these limitations, we found clear and significant group differences. Specifically, the non-retrieval time *t*_0_ was significantly higher for MCI patients (*M* = 4.64, *SD*= 1.78) than for healthy controls [*M* = 1.77, *SD* = 0.70, t(29.30) = 7.40, *p* < 0.0001]. Conversely, we found that the *F* parameter was lower in patients (*M* = 0.48, *SD* = 0.32) than in controls [*M* = 1.00, *SD* = 0.27, *t*(45.70) = –6.16, *p* < 0.0001]. The non-retrieval time parameter *t*_0_ is understood as the sum of sensorimotor processes that surround memory retrieval, including the time to properly process and recognize a stimulus (word or picture) and the time to prepare and execute a motor response. In this sense, the dramatic increase in non-retrieval time observed in MCI patients is consistent with well-established previous findings in the literature, where sensorimotor dysfunction is often associated with cognitive decline (*60*, *61*) and might foreshadow the onset of dementia (*62*). Unlike *t*_0_, the latency parameter *F* directly scales the retrieval time. Previous work (*63*) suggests it plays an analogous role to the decision boundary in accumulator models, indicating how much effort an individual can allocate to ensure a successful retrieval process. Thus, the lower value of *F* in MCI patients would indicate either reduced attentional resources or a compensatory shift of the speed-accuracy tradeoff to account for the increased non-retrieval times.

### Relationship to Behavioral Data

Throughout this paper, we have discussed the utility of model-based parameters (in this case, the speed of forgetting and its interpretable counterpart, the SGMA score) as an alternative to behavioral data. One might wonder, however, how our model parameters would compare to the behavioral data (response times and accuracies) from our test itself. In principle, such a comparison is circular: because an item’s response time and accuracy are tied to its speed of forgetting (see Equation 4) and the software uses an item’s speed of forgetting to determine when to present the next test item, both measures should be similarly connected. In fact, we found the mean test-retest reliability of response times was identical to that of SGMA scores (*r* = 0.72), while that of accuracy scores was lower but still reliable (*r* = 0.46). In addition, neither measure adds information to the SGMA score; expanding the logistic classifier to include either the mean response times or the mean accuracy for each session does not improve its discrimination accuracy (SGMA + mean RT: Accuracy = 86%; SGMA + Mean Accuracy: Accuracy = 83%).

### Limitations

Finally, our results must be understood within the context of some important limitations. The relatively small sample size of 51 participants, while sufficient to demonstrate proof of concept and establish reliability across over 2,000 testing sessions, limits the generalizability of the specific diagnostic thresholds and classification accuracies reported here. The cohort was also relatively homogeneous in terms of education level and was recruited from a single academic medical center, which may not reflect the full diversity of the aging population.

Another limitation is the sex imbalance in our sample: the higher proportion of males in the MCI group could not be fully controlled and represents, therefore, a potential confound. However, despite the prominence of the sex effect in our analyses (β = −7.61; See Table 1), the imbalance did not seem to affect the overall pattern of results. For example, the reliability of the SGMA score was comparable when computed separately for male (*r* = 0.61) and female participants (r = 0.68). Most importantly, when sex was added to SGMA as a predictor in the logistic classifier, its classification accuracy improved only marginally (from 82% to 88% for single lessons), despite the strong sex imbalance between the groups. The latter finding strongly suggests that group differences in SGMA scores are driven by the underlying differences in memory function between patients and controls, despite their group composition.

One of the advantages of SGMA is its surprising resistance to practice effects: as noted above, our practice effect amounts to only a 0.2% increase over consecutive assessments. This is perhaps consistent with previous findings that the degree to which individuals benefit from retesting is also indicative of cognitive decline and dementia (*64*, *65*). While this practice effect results in significant changes over time in our study’s weekly assessment schedule, it is likely negligible when the test is administered at more realistic intervals, such as monthly. Yet, the fact that practice effects persist (at least in healthy controls) within our adaptive assessment tool suggests a number of possible research questions. Because different stimuli were used in each testing session, practice effects in our study cannot be due to either familiarity or implicit memory of the materials, instead, they could be due to improved sensorimotor abilities (that is, greater facility at interacting with the interface) or, perhaps, to an underlying generalizable cognitive benefit from repeated retrieval practice. In the future, an improved analysis to separate the evolution of sensorimotor and cognitive parameters across sessions, as advocated above, might provide a way to address this question.

## Conclusions

This study advances the assessment of age-related memory impairment by demonstrating, for the first time, computational phenotyping as a viable alternative to traditional neuropsychological testing. Specifically, our novel assessment method, the Seattle-Groningen Memory Assessment (SGMA), demonstrated that model-based measures derived from a well-established theory of memory consolidation can achieve diagnostic accuracy comparable to standard clinical instruments while offering substantial practical advantages: remote administration, resistance to practice effects, adaptivity that reduces participant burden, and repeatability that enables high-frequency longitudinal monitoring.

SGMA’s ability to discriminate between individuals with MCI and cognitively unimpaired controls with 87% accuracy (which matches the performance of the widely used MoCA test) is particularly noteworthy, given that it accomplishes this through an 8-minute, unsupervised online task focused specifically on memory function. This performance, achieved across 53 different lesson topics and maintained in both recognition and recall modalities, demonstrates the robustness and generalizability of the underlying computational approach. The strong correlations observed across diverse materials and testing formats confirm that SGMA captures a stable, trait-like characteristic of memory consolidation rather than task-specific performance.

Beyond diagnostic classification, the longitudinal design revealed meaningful group differences in learning trajectories, with MCI participants showing attenuated practice effects over time compared to controls. This finding suggests that computational phenotyping may be sensitive not only to static differences in memory function but also to the dynamic progression of cognitive decline—a critical capability for monitoring disease progression and treatment response. The observed group-by-modality interaction, wherein MCI participants derived less benefit from the deeper processing demands of recall tasks, further illustrates how model-based approaches can distinguish encoding and consolidation deficits from retrieval difficulties, a distinction that traditional accuracy-based measures cannot make.

Finally, an important aspect of our results is the level of involvement that we were able to maintain in our cohort. Unlike most assessments, our brief online test was extremely well received by participants. This is demonstrated by two measures. The first is the remarkably low level of attrition in our sample: our participants completed, on average, 39 of the 48 weekly assessments, and none of them withdrew from the study. The second is the extraordinary level of engagement that participants displayed throughout the course of the study. In many cases, they spontaneously re-engaged with the materials, re-opening completed sessions out of personal interest in the stimuli. Forty-three participants (84%) spontaneously decided to repeat a session at least once, with each lesson being repeated at least once by one participant, with some materials (e.g., types of cheese and constellation names) being repeated up to 17 times. Because participants were not given any feedback about their performance, their re-engagement was likely driven by their intrinsic interest in the materials. We believe that such high level of engagement was made possible by two factors: (1) the fact that the SGMA score is largely independent of the specific materials used, which allowed us to generate sets of stimuli that could be relevant and interesting to our participants; and (2) its adaptive nature, which ensured that participants could perform the task with high accuracy despite their memory impairments— even MCI patients could respond correctly 83.7% of the time. Although future studies are needed to examine the limits at which our adaptive assessment can be employed, we believe that these results suggest it could be successfully applied in populations with even greater forms of cognitive impairment.

Taken together, these results have several practical implications. The current underdiagnosis of Alzheimer’s disease and related dementias stems in part from limited access to specialized neuropsychological services, particularly in rural and underserved communities. By enabling remote, self-administered assessment that requires no specialized training to conduct, is well received by patients, and is resistant to the practice effects that plague traditional instruments, computational phenotyping could substantially expand access to memory screening and monitoring. The adaptive nature of the task makes the assessment efficient and appropriate for a wide range of cognitive abilities, from intact function through mild impairment.

### Future Directions

Future research should focus on several key priorities. First, validation in larger, more diverse cohorts is essential to establish population-based norms and refine diagnostic thresholds. Second, long-term prospective studies are needed to determine whether SGMA scores can predict conversion from healthy controls to MCI or from MCI to dementia, and whether changes in SGMA scores correlate with disease progression as measured by traditional clinical and biological markers. Third, integration with biomarker data (such as neuroimaging, cerebrospinal fluid, or blood-based markers) would clarify the relationship between computational phenotypes and underlying neuropathology. Fourth, extension of this computational phenotyping approach to other cognitive domains beyond declarative memory could provide a comprehensive assessment battery suitable for differential diagnosis across various neurodegenerative conditions. Even within the limited scope of our model, the values of its ancillary parameters, such as the non-retrieval time *t*_0_ or the retrieval scale *F* (Equation 4) seem to provide important additional information that complements memory diagnostics. These parameters could play a critical role, for example, to correctly assess cognitive decline in pathologies that are more directly associated with motor impairments, such as Parkinson’s Disease, or with dysexecutive impairments, such as stroke and TBI. Thus, we believe that future studies should focus on better algorithms to estimate these parameters concurrently and independently of the SGMA score (see (*59*), for a recent promising attempt in this direction), as well as additional empirical tests with different populations (see (*34*), for an example application). Finally, although SGMA and its underlying model-based assessment have proven to be reliable across a variety of stimuli (Figure 6), the specific stimuli set used in different lessons do exhibit residual discrepancies in both absolute scores and degree of separation between groups. Because our analysis has ruled out the stimulus format (text vs. image) as a possible factor (see Table 1), future work will be needed to clarify the source of these differences and create a reliable database of well-characterized stimuli. Combining all these aspects will hopefully allow us to further reduce the overlap in MCI and healthy participants’ SGMA distributions as well as the variability across materials (Figure 7).

In conclusion, this study establishes computational phenotyping as a promising and theoretically principled approach to memory assessment that addresses many limitations of traditional neuropsychological testing. By grounding measurement in an explicit model of memory consolidation, SGMA provides interpretable, reliable, and repeatable assessments that can be administered remotely without specialized personnel. While further validation is needed, this approach has the potential to transform how we screen for, diagnose, and monitor cognitive decline in aging populations, ultimately contributing to earlier detection and more effective management of Alzheimer’s disease and related disorders.

## Data Availability

All data produced are available online at our osf repository

https://osf.io/kq5d9/

## Acknowledgements

We would like to thank Theresa Kehne and Lali Ghate from the Alzheimer’s Disease Research Center for their help in recruiting the participants, as well as all participants and student helpers for their contributions. We thank Thomas Wilschut from Precision Cognition Labs B.V. in Groningen, Netherlands for his support in the design of the recall sessions and discussions about analyses; and Alyssa Williams from Mississippi State University and Anais Capik, Sara Ulibarri, Bridget Leonard, Nevada Simpson, Lily Makaryan, and Ineeya Senthil Nathan Kayal from the University of Washington for their support in collecting data from the recall sessions.

## Data Reporting

The datasets generated and/or analyzed during the current study are available in the “Computational Phenotyping of Memory for Early, Ongoing Assessment of Mild Cognitive Impairment” repository, available through the Open Science Framework at https://doi.org/10.17605/OSF.IO/KQ5D9.

## Supplementary Figures

**Supp Fig 1.**
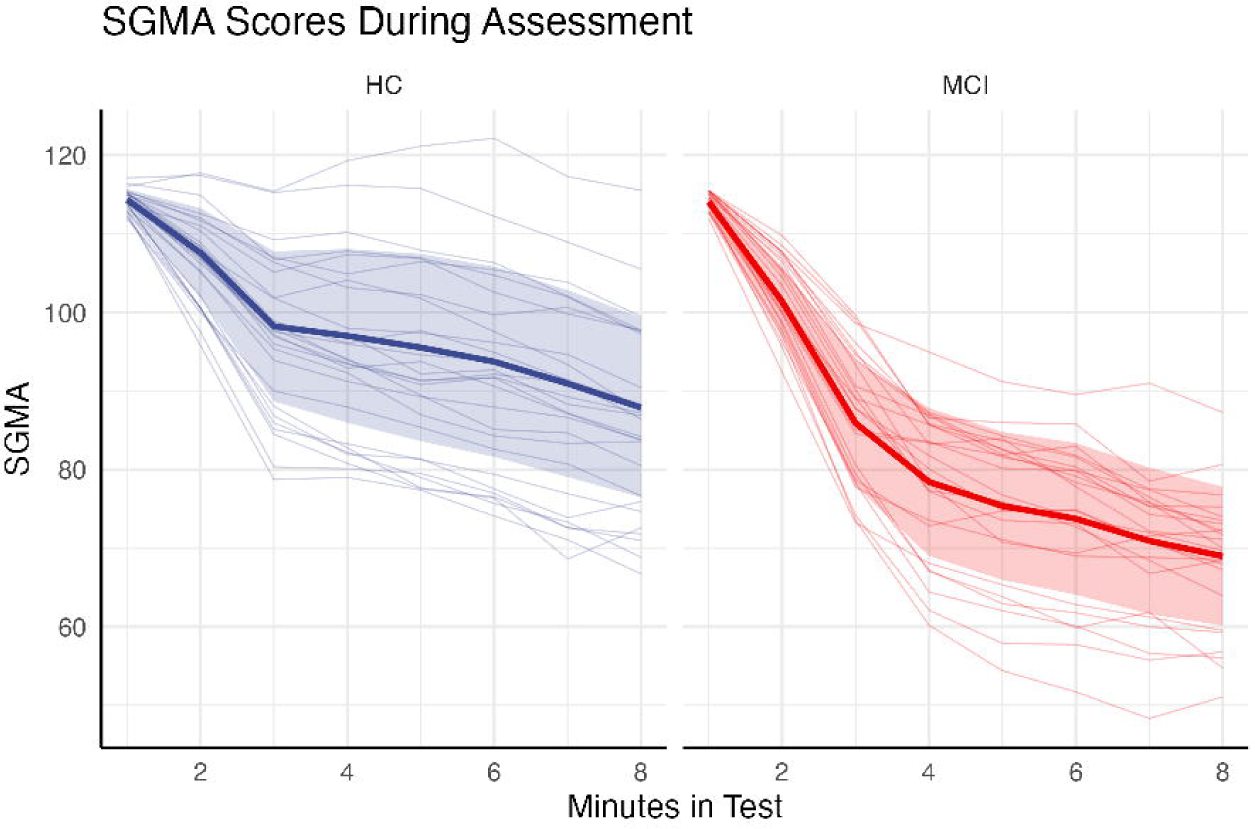
Timecourse of the SGMA score over time. SGMA values calculated as a function of the data available at different minutes in the test, averaged across all lessons within a participant and plotted separately for healthy controls (left) and MCI participants (right). Thin lines represent the mean timecourse of the SGMA score for each individual participant; thick lines and ribbons represent the group means and standard errors. Note that, within each group, the estimate passes the inflection point (the “elbow”) within the first few minutes, and the differences between groups remain stables after > 4 mins, even if the estimates keep being refined.

**Supp Fig 2.**
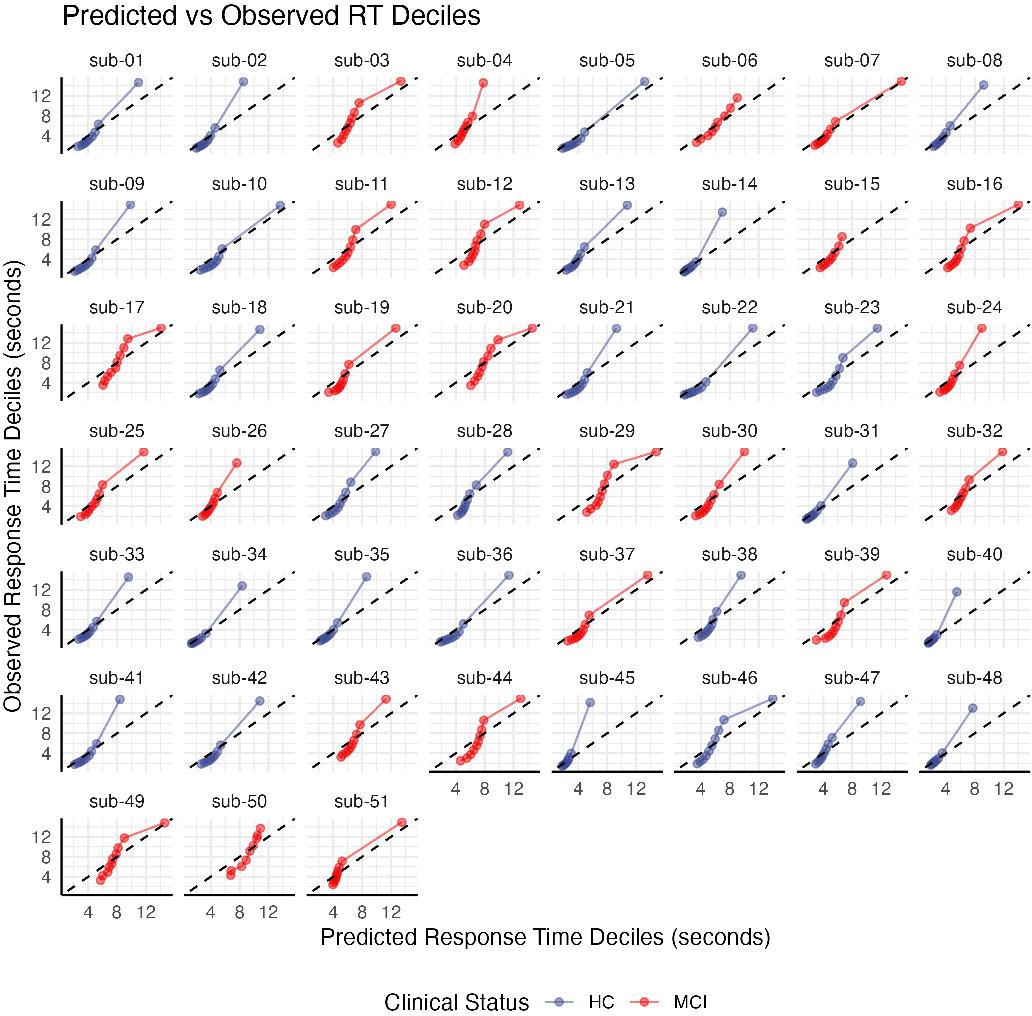
Model fit for individual response times. Decile plots, showing the predicted vs. observed RTs corresponding to the response in the 1^st^, 2^nd^, …, 10^th^ decile for each participant (note that the 10^th^ decile, not included in Fig 5, contains abnormal response outliers that are not captured by the model).

**Supp. Fig 3.**
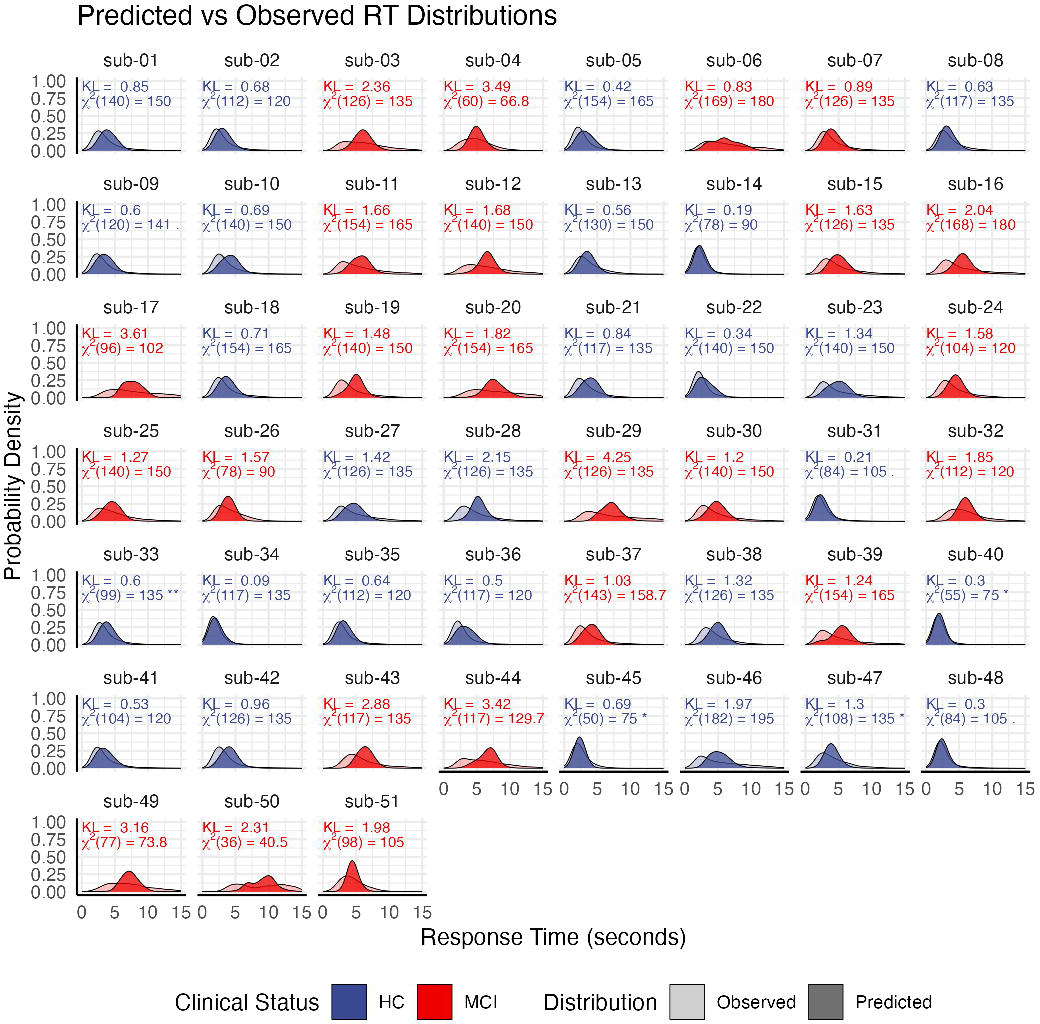
Predicted vs. Observed RT Distribution For Each Participant. Kullback-Leibler divergence and χ^2^ statistics, which provide a measure of the difference between distributions, are reported for each participant. * *p* < 0.05

**Supp. Fig 4.**
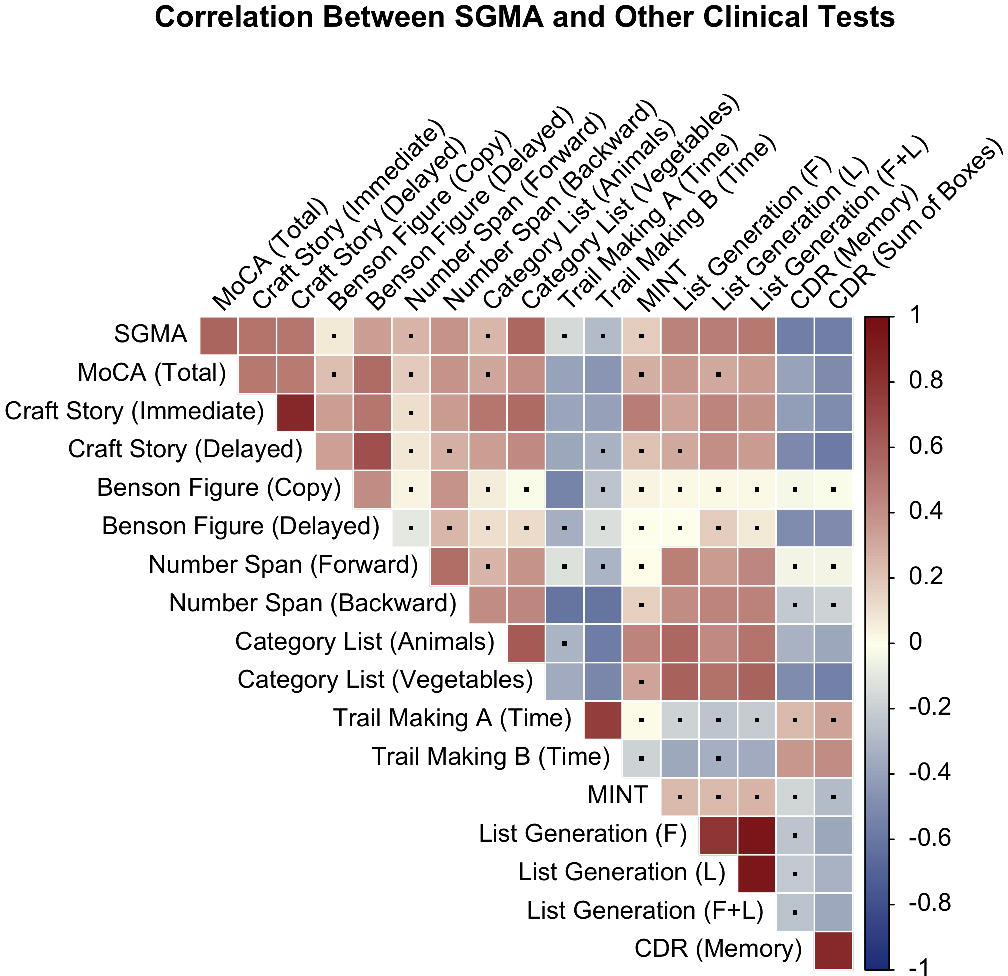
Correlation matrix between all the main neuropsychological assessments, including SGMA. Colors indicate Pearson correlations; dots represent correlations that did not reach significance (*p* > 0.05).

**Supp Fig 5.**
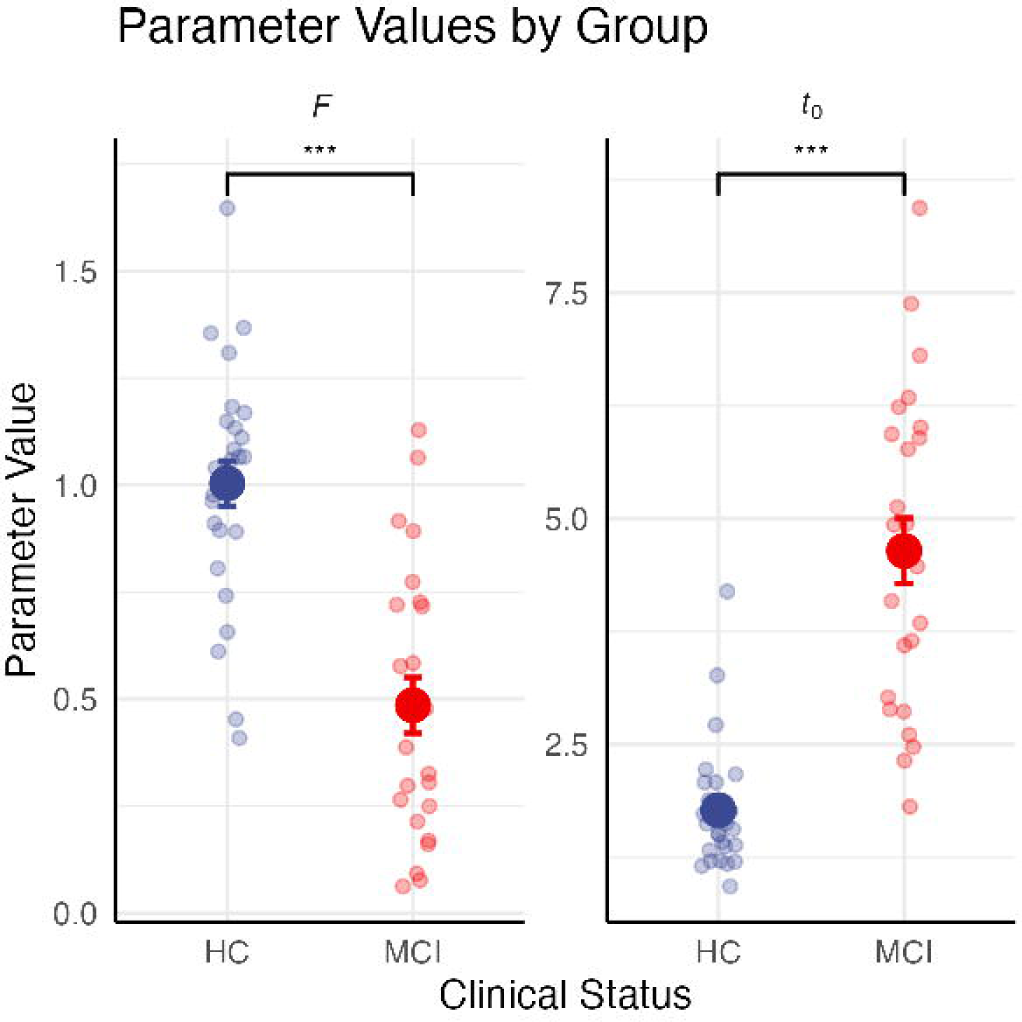
Mean values for the response time parameters *F* (left) and *t*_0_ (right) estimated for healthy controls (blue) and MCI patients (red). Small points represent estimates for each individual participant, large points represent group means, and error bars represent standard errors of the mean (*** *p* < 0.001).

